# An agent-based model of Epstein-Barr virus transmission and vaccination

**DOI:** 10.64898/2026.11.10.26367734

**Authors:** Bridgette Amoako, Zhongda Huang, Edward Thommes, Warren Hare, Monica Cojocaru

**Author notes:** former affiliation: Global Medical Affairs, Sanofi Vaccines, 1755 Steeles Ave. W, North York, ON M2R 3T4.

## Abstract

Epstein-Barr virus (EBV) infects over 90% of adults worldwide, causing infectious mononucleosis (IM) in adolescents and young adults and contributing to multiple malignancies. Despite its global burden, population-level transmission modeling of EBV remains limited. We present the first population-level agent-based model (ABM) of EBV transmission. The model is implemented with 10,000 agents and calibrated to age-specific seroprevalence data from the US National Health and Nutrition Examination Survey. Two complementary calibration methods are evaluated, each with its own distinct advantages: Approximate Bayesian Computation (ABC) and Bayesian Optimization with Expected Quantile Improvement (BO–EQI). ABC directly yields an approximate distribution of transmission parameters for propagation through vaccination scenarios. BO–EQI uses a limited simulation budget to identify well-fitting parameter vectors, particularly as the number of fitted parameters increases. We estimate parameter uncertainty at each resolution in a separate Gaussian-kernel posterior analysis, which requires additional simulations and an explicit model for discrepancies between simulated and observed prevalence. As an illustrative application, we simulate the population-level effect of hypothetical EBV vaccines over a 100-year projection horizon. Similar to a previous modeling study, our results suggest that vaccine durability is at least as critical as efficacy, and that a vaccine with direct protection against only EBV may persistently shift IM burden to older ages, an effect our longer simulation horizon reveals to be a durable equilibrium shift rather than a transient cohort phenomenon. These findings highlight the importance of careful vaccine target product profile design; in particular, our results suggest that dual efficacy against both EBV infection and IM progression represents the most promising achievable target profile, and should be a core design objective for next-generation EBV vaccines.

## 1 Introduction

Epstein–Barr virus (EBV) is one of the most widespread human pathogens, with seroprevalence exceeding 90% in adults worldwide^1,2^. Primary infection usually occurs in childhood, where it is often asymptomatic, but acquisition during adolescence or early adulthood frequently results in infectious mononucleosis (IM)^3^. Following infection, commonly through saliva^4^, EBV establishes latency within B cells and can periodically reactivate, leading to viral shedding and continued opportunities for transmission^1^. Beyond illness, EBV has been shown to result in several malignancies, including multiple sclerosis (MS), Hodgkin’s lymphoma (HL), nasopharyngeal cancer and gastric cancer^5–7^. Despite its global burden, the transmission dynamics of EBV is not as thoroughly studied as those of other common viruses like influenza. The difficulty arises from asymptomatic infections, heterogeneous shedding, and the challenge of measuring interpersonal contacts relevant for transmission.

Over the decades, attempts to control EBV infection and its sequelae have focused on managing symptoms of IM, suppressing viral replication, and preventing EBV-driven malignancies. Antiviral therapies such as acyclovir, ganciclovir, and valganciclovir have been trialed, primarily targeting lytic replication^8–10^. Lytic replication, or the lytic cycle, is a viral reproduction process where a virus infects a host cell, replicates its genetic material, and ultimately destroys the host cell to release new viral particles. While these drugs can reduce viral shedding in the short term, clinical studies have shown little benefit in altering the course of infectious mononucleosis or preventing latency, since EBV persists as an episome within B cells^11^. Similarly, corticosteroids have been used to manage severe IM complications such as airway obstruction, but they are not recommended for routine treatment due to limited benefit and potential risks^12^. These results highlight the challenge of intervening after primary infection has occurred.

Vaccination against EBV has long been considered an attractive preventive strategy, particularly given its contribution to both acute morbidity and long-term cancer risk. The most advanced vaccine candidate to date was a recombinant glycoprotein (gp) 350 subunit vaccine developed by GlaxoSmithKline Biologicals (Rixensart, Belgium), which was tested in a large phase 2 trial. This vaccine reduced the incidence of IM by 78% (95% CI [1.0% - 96.0%]) but did not prevent EBV infection itself^13^. More recently, efforts have shifted toward vaccines targeting multiple viral antigens, including those involved in viral entry into epithelial and B cells^14^. Advances in nanoparticle platforms and mRNA technologies have renewed interest in EBV vaccine development, with several candidates currently in preclinical or early clinical stages^15,16^. These approaches aim not only to reduce IM but also to mitigate downstream risks of EBV-associated cancers and autoimmune diseases.

Mathematical modeling offers a valuable framework, allowing exploration of how age structure, infection outcomes, and behavioral factors shape population-level patterns of EBV spread. Such models also provide tools to test hypothetical scenarios such as vaccination programs, which are increasingly under consideration given the morbidity associated with IM and EBV’s role in cancer risk. Previous research on EBV transmission modeling has employed diverse approaches to understand infection dynamics and inform public health strategies. Goscé et al. (2019)^17^ represents the most comprehensive population-level mathematical model of EBV transmission to date, explicitly incorporating age structure and vaccination scenarios. Calibrated to seroprevalence data from England, the compartmental model showed that vaccine duration of protection has a greater impact on EBV and infectious mononucleosis (IM) prevalence than efficacy. Short-lived immunity deferred infection into adolescence, where IM risk is highest, while longer-lasting protection reduced both transmission and disease burden. The study also noted that vaccinating younger age groups could unintentionally delay infection to later ages, thereby increasing the likelihood of IM.

Exploring infection dynamics in a different way, Shapiro et al. (2008)^18^ created an agent-based simulation of EBV infection within a host, enabling exploration of infection parameters. The agents of this model are EBV virus, B cells and T lymphocytes. Addressing age-related risk directly, Huynh and Adler (2012)^19^ applied an age-dependent framework to explain why infectious mononucleosis (IM) peaks in adolescence, focusing on in host dynamics rather than interventions. A joint statistical model linking Danish seroprevalence with IM incidence was employed by Rostgaard et al. (2019)^20^ to estimate age- and sex-specific infection hazards and attack rates. In Balfour et al. (2013)^3^, U.S. NHANES (The National Health and Nutrition Examination Surveys) data is used to estimate the age-specific prevalence of EBV infection in individuals aged 6–19 years. It identifies demographic and socioeconomic factors associated with earlier EBV acquisition, providing updated population-level insights into infection timing in the United States. The agent based model we develop also uses this data.

In this study, we develop a flexible, long-horizon agent-based model of EBV transmission and IM development. It is the first agent-based model, and only the second population-level transmission model^17^ of EBV. The first model, a compartmental model, was parametrized using data from England. The agent-based model we develop is calibrated with data from the United States. An agent-based model was chosen over a compartmental one in order to facilitate the simulation of heterogeneous populations, and to allow the tracking of complex individual health state histories (e.g. age at EBV infection and IM onset) without requiring a combinatorial explosion in the number of compartments. Being able to resolve these histories in detail is critical to the future addition of EBV-associated sequelae, e.g. MS. We also use this model as a testbed for comparing two calibration strategies for stochastic agent-based models: Approximate Bayesian Computation (ABC) and Bayesian Optimization with Expected Quantile Improvement (BO-EQI). We distinguish point calibration from uncertainty quantification. ABC directly supplies a joint parameter sample for the vaccination projections. BO–EQI directs simulations toward finding a well-fitting point under a fixed evaluation budget. We additionally examine parameter uncertainty through a separate importance-sampled posterior; its intervals are not an output of BO–EQI itself. Finally, we use the calibrated model to examine illustrative hypothetical vaccine scenarios selected to be comparable with previous EBV modeling work.

### 1.1 Paper Structure

The remainder of this paper is organized as follows. Section 2 describes the agent-based model framework. Sections 3 and 4 present two methods of calibration for the estimation of the probability of transmission per meaningful contact in the agent based model of EBV infection: the Approximate Bayesian Calibration (ABC) method and Bayesian Optimization with Expected Quantile Improvement (BO–EQI) algorithm. Section 5 presents the outcomes observed when vaccination against EBV is introduced, for different target product profiles. Finally, Section 6 concludes the paper and outlines directions for future research.

## 2. Methodology

### 2.1 The agent-based model and synthetic population

We developed an agent-based model implemented in R to study the transmission of Epstein–Barr Virus (EBV) and the occurrence of Infectious Mononucleosis (IM) within a synthetic population. At initialization, the model generates a population of agents distributed across age and sex-assigned-at-birth to reflect the demographic structure of the target setting, with some agents seeded as EBV-infected. The demographic structure of the model is informed by population birth rates^21^ and age-stratified mortality rates^22^. Social mixing patterns are defined by an age-structured social contact matrix of the USA^23^, as implemented in the Bernadette R package^24^.

Transmission-related parameters capture the risk of EBV infection and of developing IM at the time of infection. The probability of EBV transmission per infectious contact is calibrated using as the target the age-specific prevalence of EBV within the US population^3^, estimated using data from the US National Health and Nutrition Examination (NHANES) survey. Calibration is performed using two methods: Approximate Bayesian Computation (ABC) and Bayesian Optimization (BO). Further details are provided in Section 3. The probability that a new EBV infection leads to Infectious Mononucleosis is also stratified by age^20^. Where vaccination is available, the information provided includes the age groups eligible for vaccination, the annual probability of vaccination in those groups, and the efficacy of the vaccine. In the base case, vaccination is not implemented. Hypothetical vaccine strategies are explored in our analysis (see Section 5 below).

A simulated scenario starts with an initial population (here, 10,000 agents distributed according to present-day US demography), a start date, and a duration. Agent attributes are summarized in Table 1 and model parameters in Table 2. Time advances in discrete steps; through experimentation, we found that a step size of 28 days adequately resolves the (relatively slow) EBV transmission dynamics. At each step, agents undergo deterministic changes (aging, waning of protection if vaccinated), and depending on their current state, may probabilistically undergo transitions to vaccinated, infected (EBV or EBV+IM), or death. This structure allows the simulation to capture how demographic processes, infection risk, and vaccination strategies shape EBV prevalence and IM incidence over time. The social contact matrix governs exposure opportunities by defining the frequency of interactions between age groups, which drives the probability of transmission. Upon exposure, susceptible individuals may acquire EBV infection. If infection occurs, an age-dependent probability governs whether the agent develops IM. Thus, an agent’s health state can be (i) EBV-seronegative; (ii) EBV-seropositive without IM at primary infection; or (iii) EBV-seropositive with IM at primary infection.

**Table 1.** Agent Attributes and Descriptions.

| Attribute | Description | Values |
|---|---|---|
| Unique identifier | Unique numeric identifier for each agent | Integers ≥ 1 |
| Age | Agent’s age in years | Continuous values ≥ 0 |
| Sex-at-birth | Agent’s sex-at-birth | Male / Female |
| EBV infection status | Indicates whether the agent is infected with EBV | True / False |
| IM infection history | Indicates whether the agent has ever had IM | True / False |
| Vaccination status | The current vaccination status of the agent | True / False |
| Survival status | Indicates whether the agent is still alive | Alive / Dead |

**Table 2.**
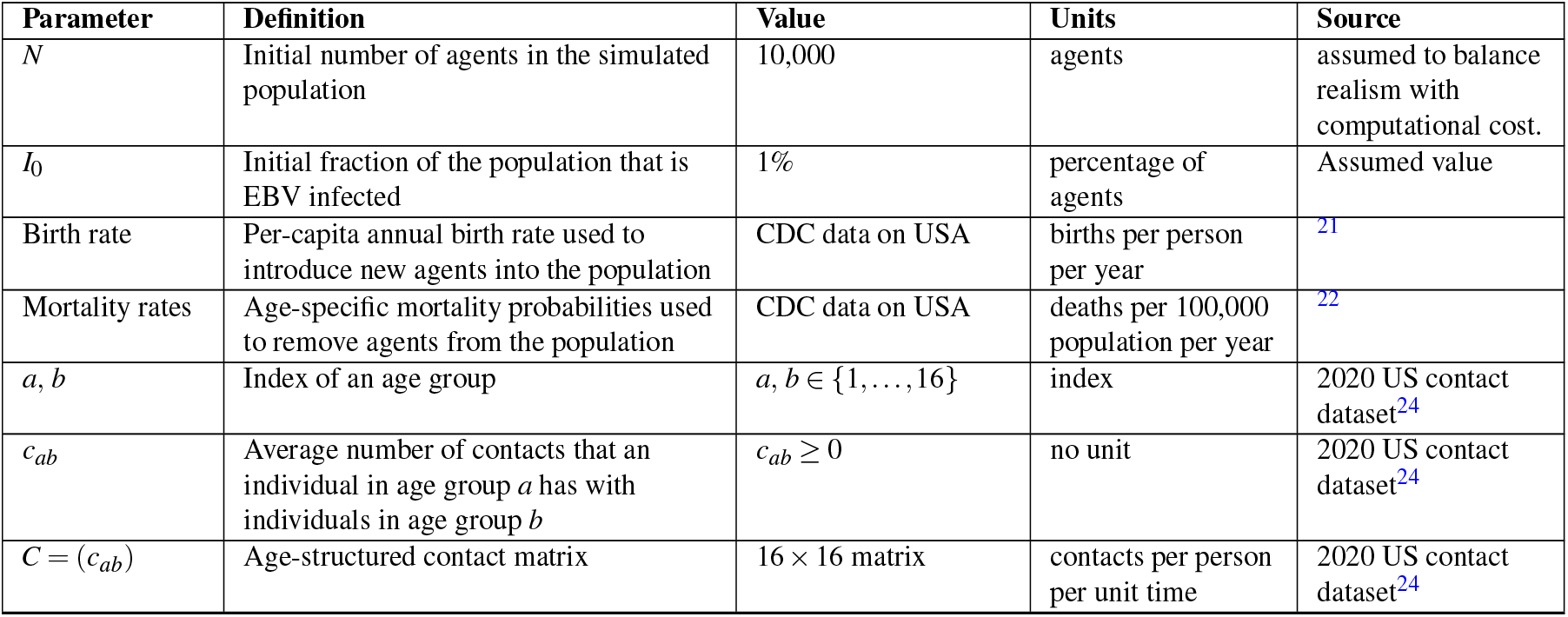
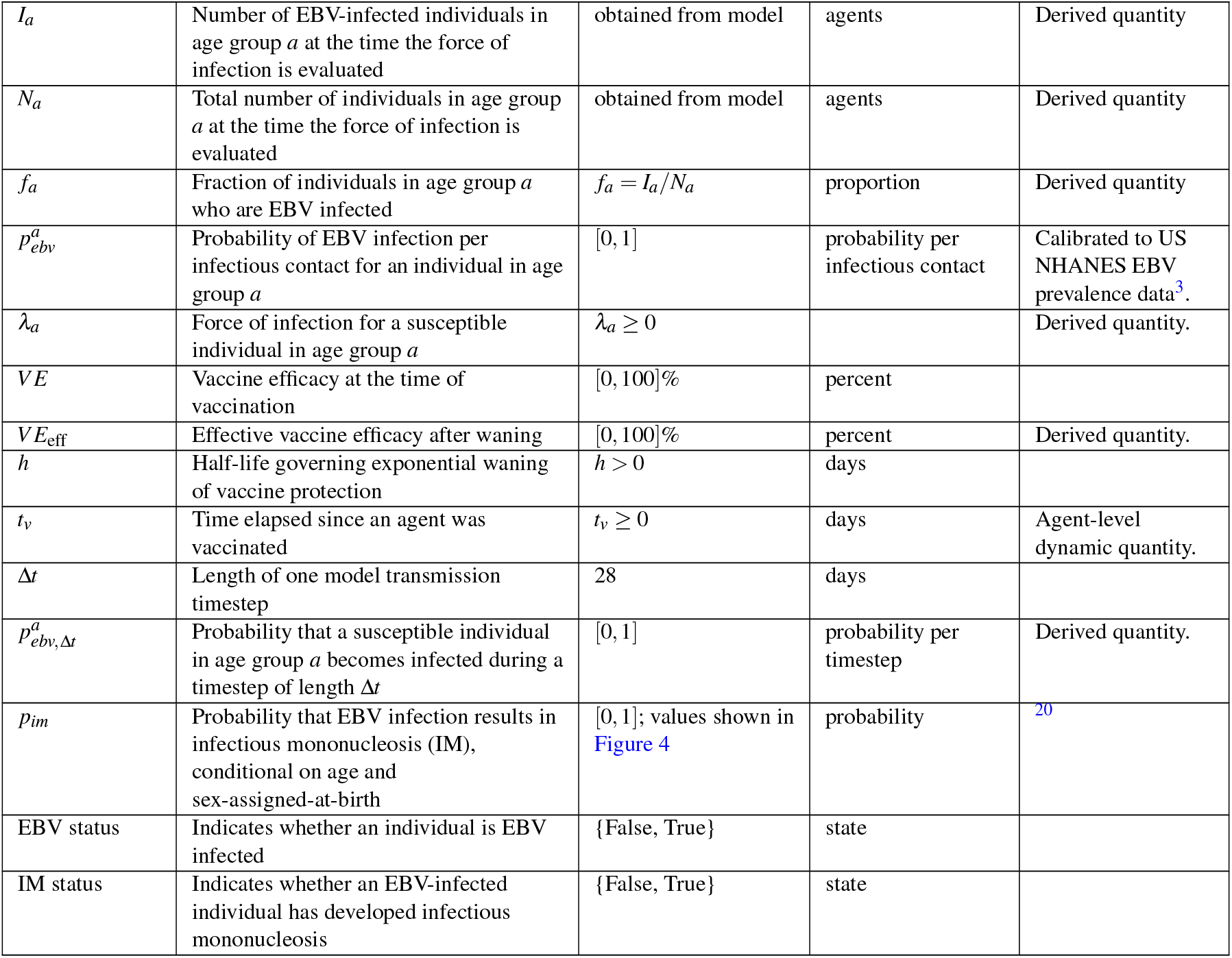
Model parameters.

| Parameter | Definition | Value | Units | Source |
|---|---|---|---|---|
| *N* | Initial number of agents in the simulated population | 10,000 | agents | assumed to balance realism with computational cost. |
| *I*<sub>0</sub> | Initial fraction of the population that is EBV infected | 1% | percentage of agents | Assumed value |
| Birth rate | Per-capita annual birth rate used to introduce new agents into the population | CDC data on USA | births per person per year | <sup>21</sup> |
| Mortality rates | Age-specific mortality probabilities used to remove agents from the population | CDC data on USA | deaths per 100,000 population per year | <sup>22</sup> |
| *a*, *b* | Index of an age group | *a*, *b* ∈ {1, …, 16} | index | 2020 US contact dataset<sup>24</sup> |
| *c*<sub>*ab*</sub> | Average number of contacts that an individual in age group *a* has with individuals in age group *b* | *c*<sub>*ab*</sub> ≥ 0 | no unit | 2020 US contact dataset<sup>24</sup> |
| *C* = (*c*<sub>*ab*</sub>) | Age-structured contact matrix | 16 × 16 matrix | contacts per person per unit time | 2020 US contact dataset<sup>24</sup> |
| *I*<sub>*a*</sub> | Number of EBV-infected individuals in age group *a* at the time the force of infection is evaluated | obtained from model | agents | Derived quantity |
| *N*<sub>*a*</sub> | Total number of individuals in age group *a* at the time the force of infection is evaluated | obtained from model | agents | Derived quantity |
| *f*<sub>*a*</sub> | Fraction of individuals in age group *a* who are EBV infected | *f*<sub>*a*</sub> = *I*<sub>*a*</sub>/*N*<sub>*a*</sub> | proportion | Derived quantity |
| *p*<sup>*a*</sup><sub>*ebv*</sub> | Probability of EBV infection per infectious contact for an individual in age group *a* | [0, 1] | probability per infectious contact | Calibrated to US NHANES EBV prevalence data<sup>3</sup>. |
| *λ*<sub>*a*</sub> | Force of infection for a susceptible individual in age group *a* | *λ*<sub>*a*</sub> ≥ 0 |  | Derived quantity. |
| *VE* | Vaccine efficacy at the time of vaccination | [0, 100]% | percent |  |
| *VE*<sub>eff</sub> | Effective vaccine efficacy after waning | [0, 100]% | percent | Derived quantity. |
| *h* | Half-life governing exponential waning of vaccine protection | *h* > 0 | days |  |
| *t*<sub>*v*</sub> | Time elapsed since an agent was vaccinated | *t*<sub>*v*</sub> ≥ 0 | days | Agent-level dynamic quantity. |
| Δ*t* | Length of one model transmission timestep | 28 | days |  |
| *p*<sup>*a*</sup><sub>*ebv*,Δ*t*</sub> | Probability that a susceptible individual in age group *a* becomes infected during a timestep of length Δ*t* | [0, 1] | probability per timestep | Derived quantity. |
| *p*<sub>*im*</sub> | Probability that EBV infection results in infectious mononucleosis (IM), conditional on age and sex-assigned-at-birth | [0, 1]; values shown in Figure 4 | probability | <sup>20</sup> |
| EBV status | Indicates whether an individual is EBV infected | {False, True} | state |  |
| IM status | Indicates whether an EBV-infected individual has developed infectious mononucleosis | {False, True} | state |  |

The model can optionally be set to demographically rebalance the population at specified intervals. To minimize any impact of this procedure on transmission dynamics, the infection status of newly introduced agents is sampled from the current age-specific EBV prevalence, thus preserving the age-specific proportion of infectious individuals. This demographic rebalance is done to maintain the initial population structure as the model is simulated to reach a stable epidemiological state before analyses. The simulation period during which the model reaches a stable state is known as the warm-up period. In Figure 3 the warm-up period is 100 years for a population of 10,000 agents, with a demographic rebalance every quarter, and the probability of EBV infection per meaningful contact, which we calibrate in Sections 3 and 4, set to arbitrary values of 4.0 *×* 10^*−*5^, 1.25 *×* 10^*−*6^, and 4.5 *×* 10^*−*5^ for ages 0-5, 6-14, and 15+, respectively.

### 2.2 Implementation of disease transmission

The probability of infection for each agent is derived from the force of infection and the daily infection probability. The model first computes the proportion of infectious (EBV-positive) individuals within each age group. As shown in Figure 1, the population is divided into 16 age groups. This proportion is expressed as 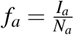, where *I*_*a*_ denotes the number of infected individuals in age group *a* and *N*_*a*_ is the total number of individuals in that age group. The vector of infectious fractions, **f** = (*f*_1_, *f*_2_, …, *f*_16_), is multiplied by the contact matrix *C* = (*c*_*ab*_), where *c*_*ab*_ represents the number of contacts a person in group *a* has with individuals in group *b*. For each age group *a*, the product 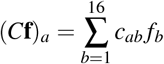 gives the number of contacts a person in group *a* makes with infectious individuals. Given the probability of infection per infectious contact 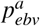, the force of infection is evaluated as

**Figure 1.**
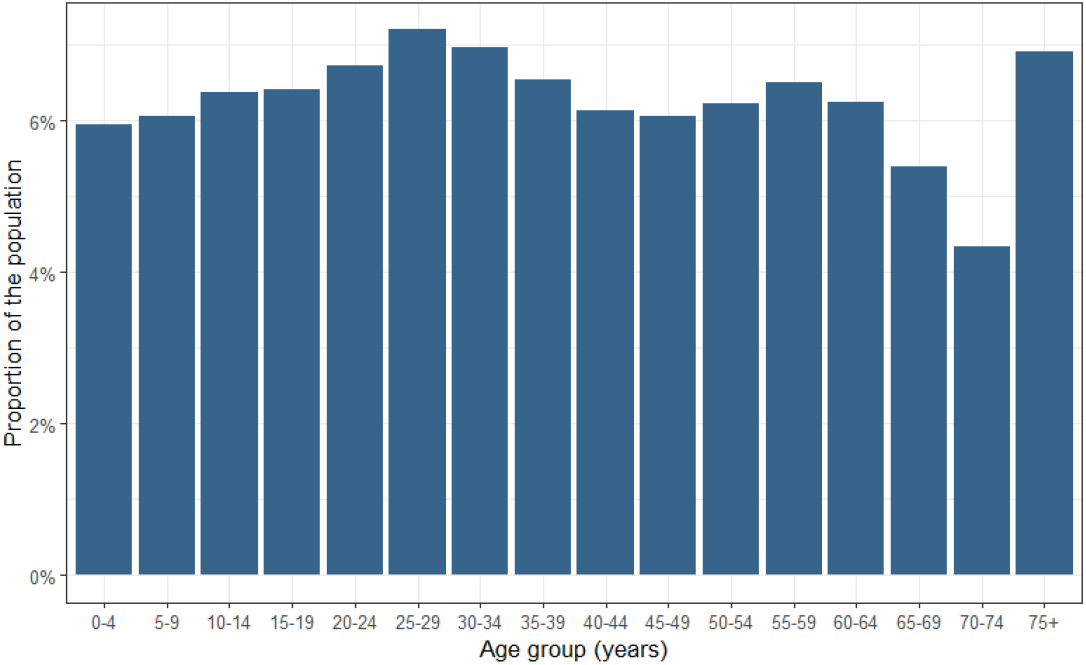
Population age distribution across 16 age groups

**Figure 2.**
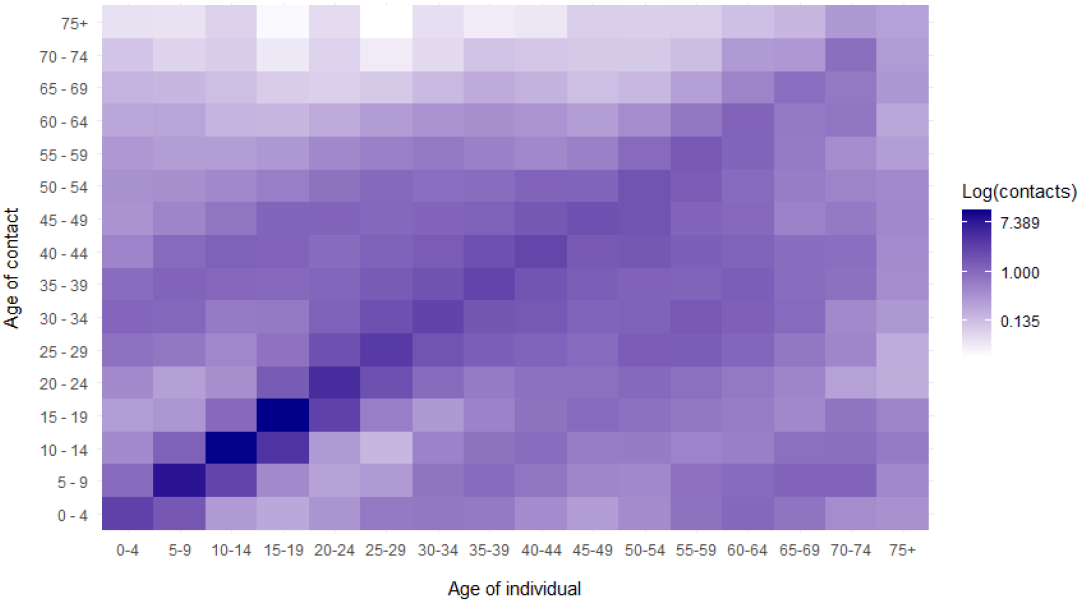
Log-scaled matrix of daily contacts per person.

**Figure 3.**
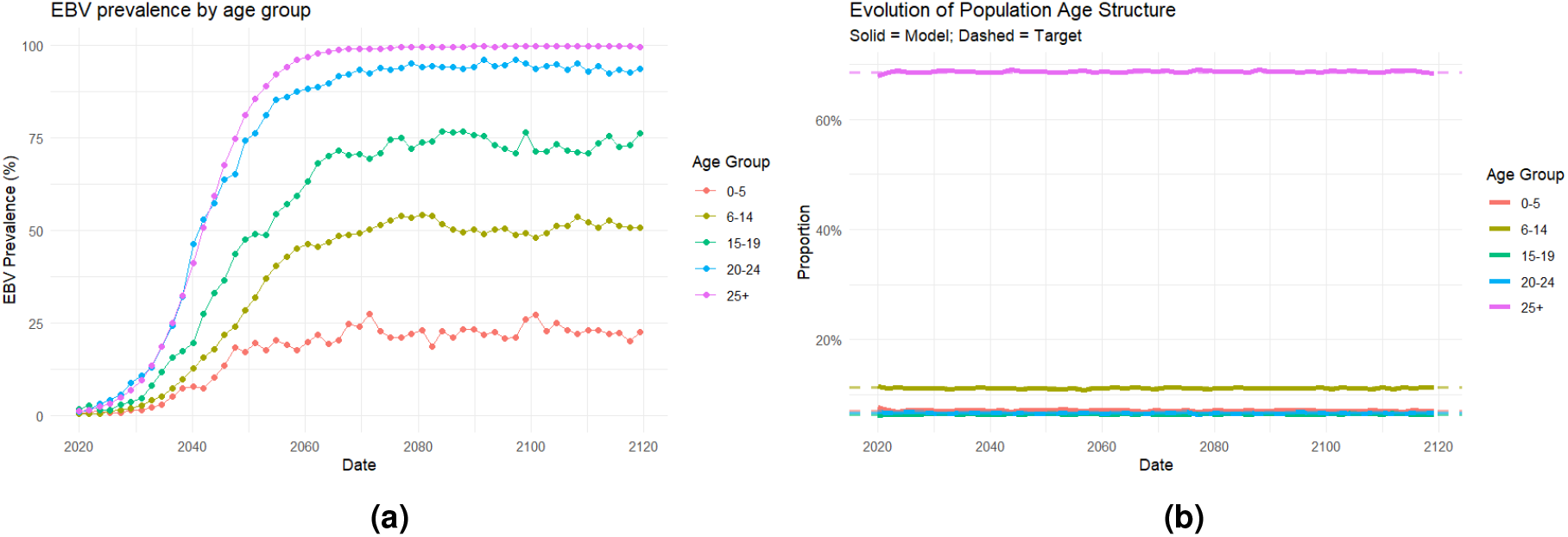
(a) For a simulation with a population of 10,000 agents, EBV prevalence reaches an endemic equilibrium state within the 100 year period; (b) Demographic rebalancing keeps the population proportions stable over the warm up period.

**Figure 4.**
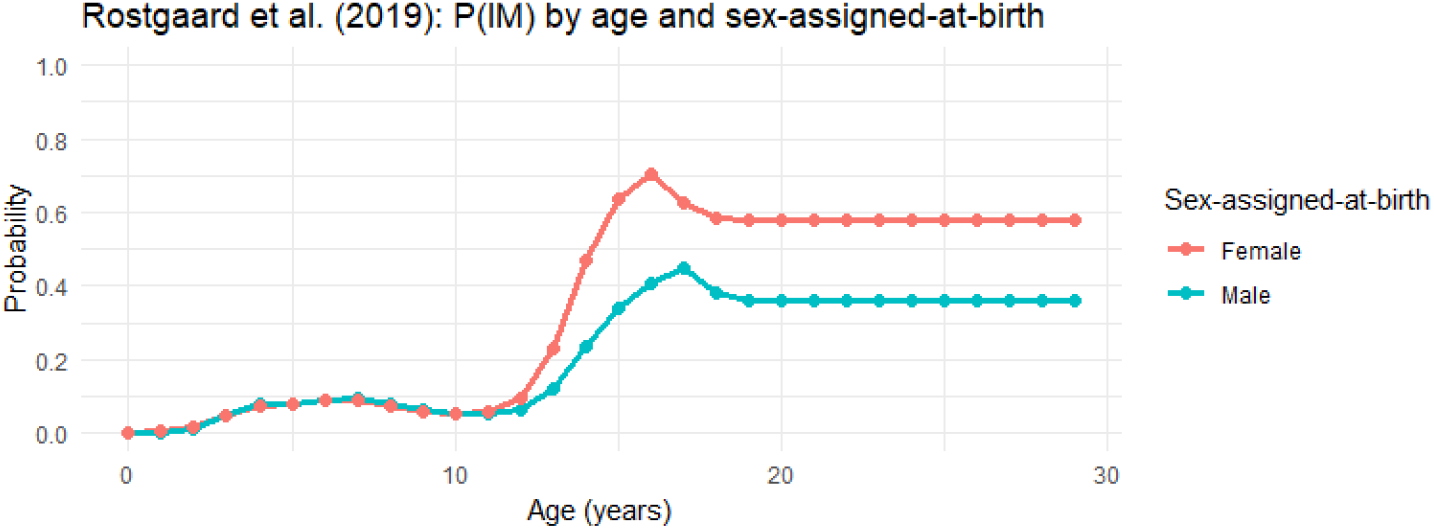
Probability of the onset of infectious mononucleosis by age and sex-assigned-at-birth; see Table 1 and Figure 1D of Rostgaard et al^20^

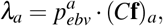

This represents the per day infection hazard for the susceptible agents in age group *a*. If the agent is vaccinated, the force of infection is reduced by a vaccine efficacy term. The model accounts for the waning of protection over time using an exponential decay function characterized by a half-life, *h*. With efficacy *VE* (in percent), and days since vaccination *t*_*v*_, the effective efficacy, *VE*_*e f f*_, is

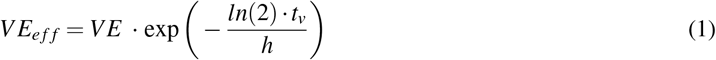

The adjusted force of infection is

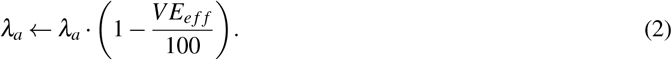

Over a timestep of length Δ*t* days,

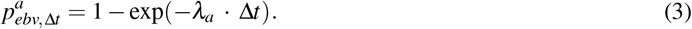

When a susceptible agent is infected, their EBV status is updated accordingly. Among the newly infected, the age-specific probability of developing IM (*p*_*im*_) determines whether they simultaneously develop IM, in which case their IM status is set to “True”.

## 3 Probability of EBV infection per meaningful contact estimates

In this section and the next (Section 4), we highlight two methods of calibrating the probability of an agent to be infected with EBV, *p*_*ebv*_, to reproduce the observed US epidemiology. Due to the challenges in tracking new infections, as they are often asymptomatic and not reported, the exact infection probability values are not available in literature. We calibrate to the US NHANES prevalence data provided in Balfour et. al (2013)^3^. The paper specifies point estimates and a 95% confidence interval for the age groups (6 *−*8, 9 *−*11, 12 *−*14, 15*−* 17, 18 *−*19). The two methods we implement are Approximate Bayesian Computation (ABC) and Expected Quantile Improvement, a form of Bayesian Optimization. The matched-budget comparison in Section 4 evaluates how well each method finds a fitted point. The posterior analysis reported there is a separate calculation and is not included in the 700-evaluation budget of either method.

### 3.1 Approximate Bayesian Computation

ABC is a likelihood-free method for Bayesian inference. In cases where the likelihood function is too complicated to formulate, ABC enables us to approximate the parameter of interest through multiple simulations. Parameters are sampled from the prior distribution, and for each draw, simulated data is generated. Both simulated and observed data are transformed into summary statistics for comparison by some distance measure. We implement the ABC-Rejection method, taking into consideration the confidence interval of the observation. The method of Latin Hypercube Sampling (LHS) is applied to generate parameters *θ*_*i j*_ within specified prior ranges, where 1 ≤ *i* ≤ *m* is the size of the prior set and *j* = 1, …, *n* is the number of age groups.

The calibration age groups used to define these parameters are (0 *−*5, 6 *−*14, 15+), which are distinct from the five age groups used to summarize model prevalence for comparison with the Balfour et al. data. This grouping reflects an assumption that individuals within these broader age ranges have similar patterns of close social contact and saliva exposure, such as contact through caregiving, shared objects, or intimate interactions, which influence EBV transmission. Broader and finer age stratifications were considered, but the three-group specification was selected as a practical balance between capturing age-related differences in transmission and limiting the number of age-specific transmission probabilities estimated from relatively sparse age-specific prevalence data. In section 4, a sensitivity experiment is conducted using the more scalable Expected Quantile Improvement method, with the number of age-specific transmission parameters varied from one to seven. Model outputs are consistently summarized as age-specific prevalence for the five age groups reported by Balfour et al.

To generate results from the *m× n* parameter matrix, we complete simulations that cover a 100 year period. For each scenario the mean prevalence within the age groups (6 *−*8, 9 *−*11, 12 *−*14, 15*−* 17, 18 *−*19) are calculated as our summary statistics, to be compared to the intervals of prevalence for matching age^3^. The running of prior simulations is perfectly parallelizable since there is zero communication needed between simulations. Hence, performance scales linearly with the number of processors. The simulations were completed on a single workstation equipped with a 12th Gen Intel Core i5-12500 CPU, with 6 physical cores and 12 logical processors, and 16 GB RAM. Using this hardware, 10,000 prior simulations were completed in approximately four hours.

We want our calibration to be consistent with not just the point estimates but also the confidence intervals of our target data. To begin with, because our summary statistics consist of probabilities bounded between 0 and 1, they are transformed using the logit function

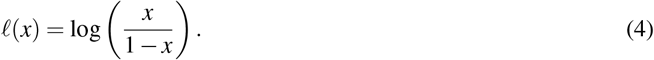

The confidence intervals of observed summaries are used to estimate standard errors

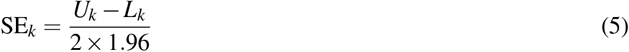

for each statistic *k*, where (*L*_*k*_,*U*_*k*_) are the lower and upper confidence limits respectively. These values are then used to scale each summary, allowing comparisons to account for uncertainty in the observed data. This is done by the equation

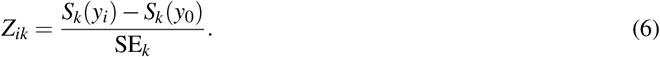

The transformed and standardized summaries are compared using the Euclidean distance,

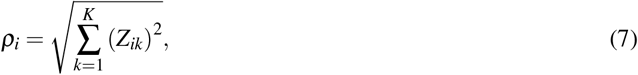

which measures how far each simulation lies from the observed summaries relative to their uncertainty. Smaller values of *ρ*_*i*_ indicate simulations that more closely reproduce the empirical data.

A series of tolerance levels *α* ∈ [10^*−*4^, 0.20] on a uniform interval with step 10^*−*4^ are examined, each corresponding to a distance threshold *ε*_*α*_, defined as the *α−* quantile of the distance distribution that determines the proportion of best-fitting simulations to retain. For each tolerance level, the retained subset of simulations is evaluated to ensure proper calibration. Standardized differences between simulated and observed summaries should have approximately zero mean, unit variance, and 95% coverage under a standard normal reference. The tolerance *α*^*\**^ that minimizes deviation from these targets is selected as optimal. The corresponding set of accepted parameters forms our posterior sample

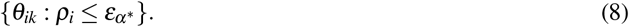

Algorithm 1 provides a pseudo-code of the ABC calibration process.

#### Algorithm 1 ABC calibration (summary, parallelized)

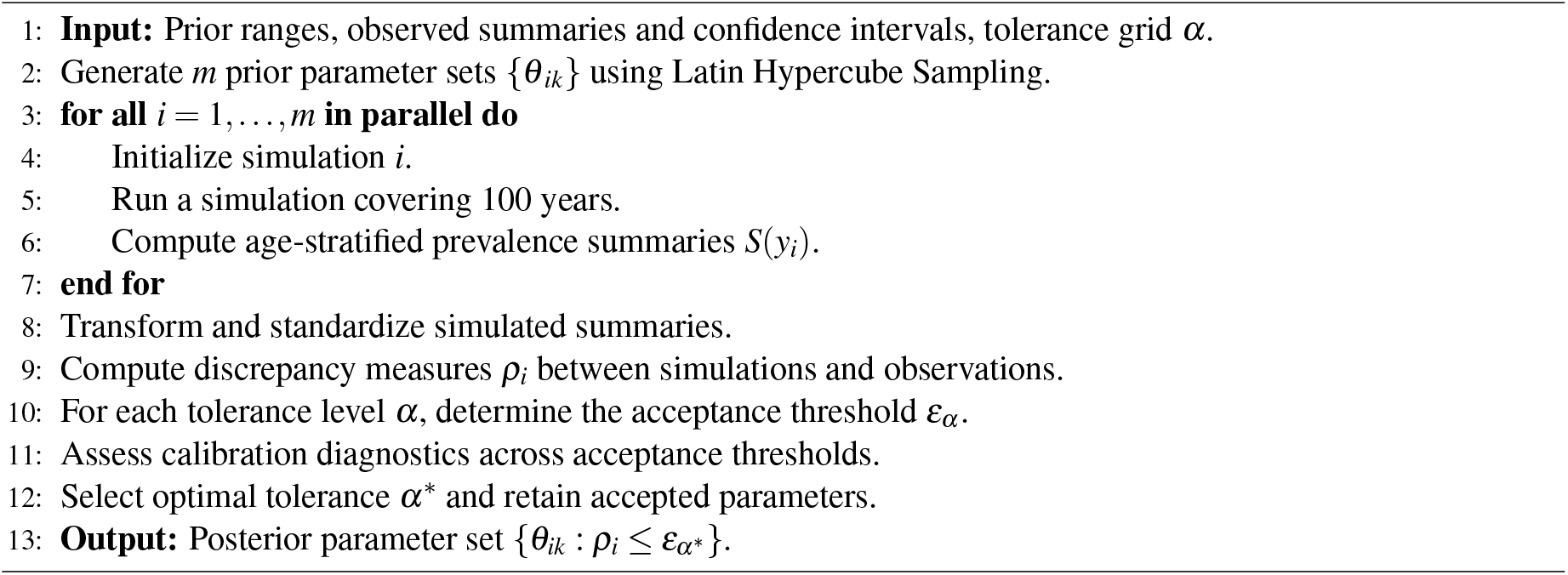

### 3.2 Discussion of Constraints on the Number of Agents and Simulations

We execute the ABC-Rejection method discussed in the previous section for different prior sample size (simulations) and the number of agents initialized, to investigate their impact on the posterior. From Figure 5, we observe a marginal return for larger number of simulations and agents. The posterior for 10,000 simulations and agents is used in our analysis. The distribution of values for each parameter for the selected number of agents and simulations (10,000) is shown in Figure 6. In Figure 7 we compare the posterior predictive to the target of observed intervals.

**Figure 5.**
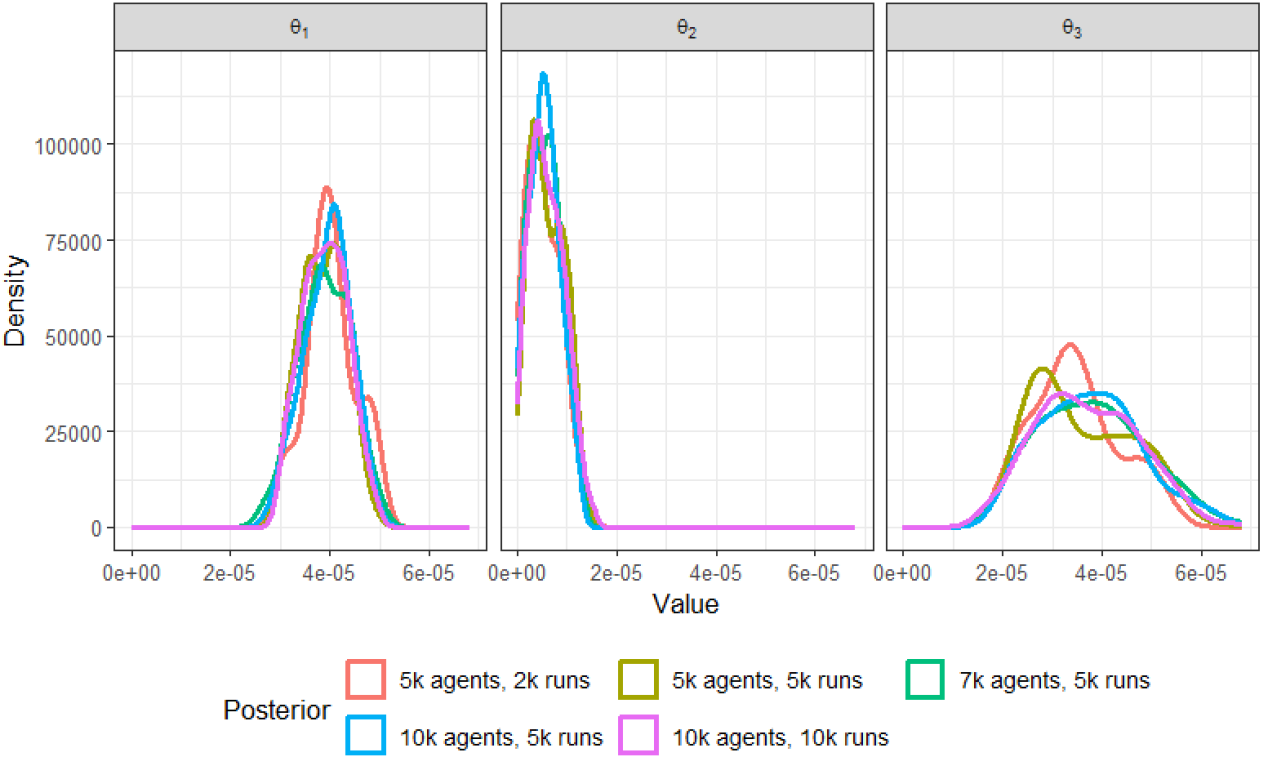
Posterior density for varying number of prior samples and the initial number of agents.

**Figure 6.**
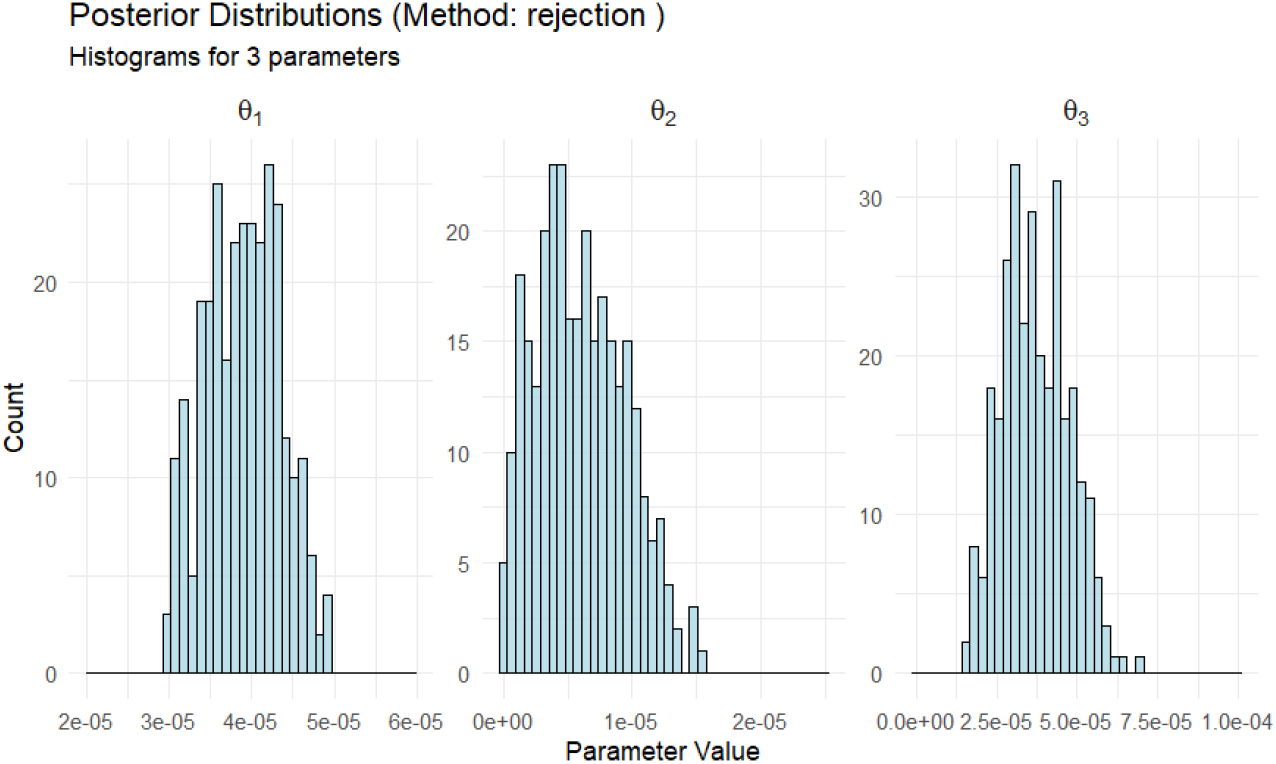
Histogram of posterior distribution of *θ*_*ik*_ (10,000 agents and 10,000 simulation runs.)

**Figure 7.**
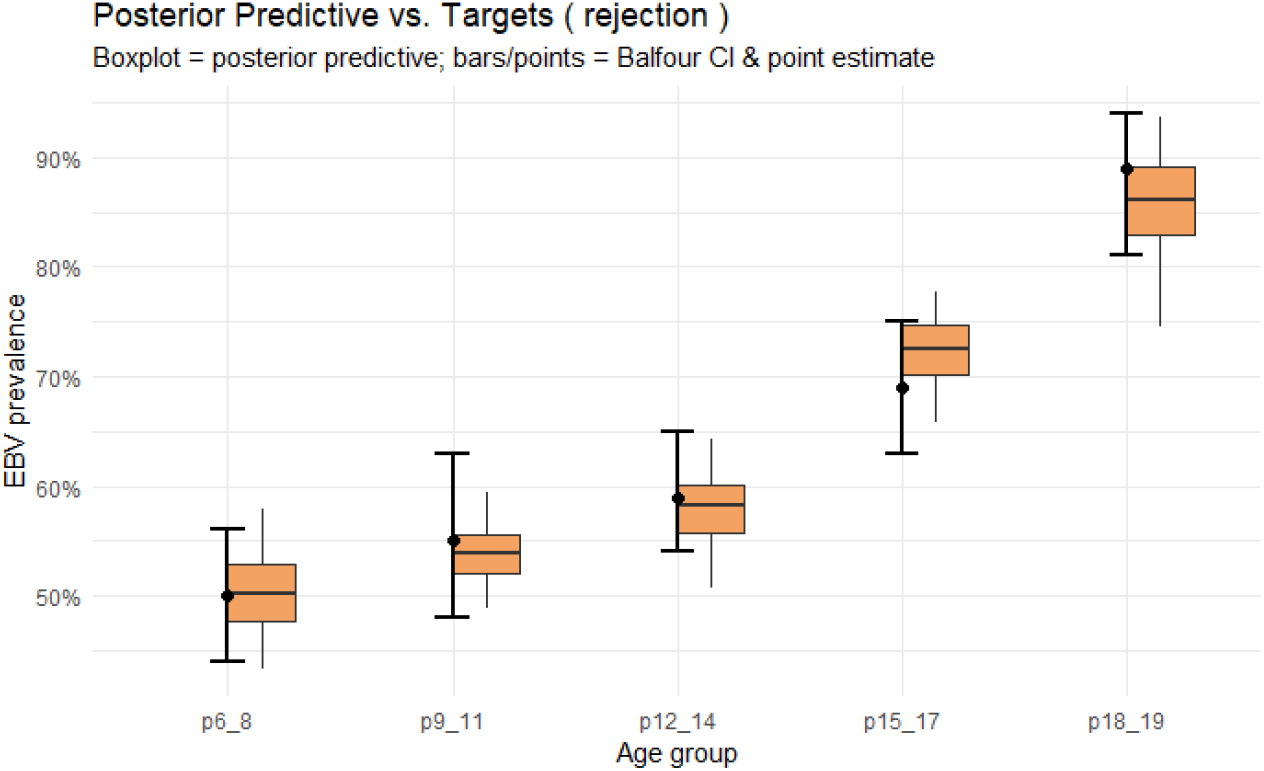
Comparison of ABM-calibrated age-specific prevalence across posterior set, with 10,000 agents (boxes showing 50% and 95% CIs) to target prevalence (error bars showing 95% CIs).

## 4 Bayesian Optimization for Calibration

### 4.1 Expected Quantile Improvement

Bayesian optimization (BO) has emerged in recent years as a powerful method for solving black-box optimization problems where each evaluation is expensive^25^. BO algorithms construct a Gaussian Process (GP) model of the objective function and use an acquisition function to decide which parameters to evaluate next. Notably, BO is well-suited for moderate-dimensional calibration problems, typically involving up to a few dozen parameters. Recent studies demonstrate BO’s practicality for ABM calibration at scale. Reiker et al.^26^ describe calibrating an agent-based malaria model as a multi-objective optimization problem with 23 parameters. By using emulator-assisted Bayesian global optimization, they could navigate the 23-parameter space and fit multiple epidemiological objectives simultaneously.

One key challenge in calibrating ABMs is their stochastic behavior: the model produces different outcomes even with the same parameter values, due to random seed variation or probabilistic rules in agent interactions. Our approach addresses this by using a Bayesian optimization framework tailored for noisy outputs. In particular, we employ an acquisition function known as Expected Quantile Improvement (EQI)^27^. The EQI criterion is a variant of the widely used Expected Improvement (EI) acquisition, modified to handle uncertainty in the output. EQI uses a GP predictive quantile to evaluate the expected improvement from an additional noisy simulation. Here the quantile describes uncertainty in the GP prediction of discrepancy. It does not directly describe the distribution of repeated ABM outcomes or provide uncertainty intervals for the infection parameters.

### 4.2 Optimization Problem Formulation

Let *θ* = (*θ*_1_, *θ*_2_, …, *θ*_*d*_)^⊤^ ∈ ℝ^*d*^ denote the vector of per-contact infection probabilities for *d* parameter age groups in the agent-based model (ABM). Each *θ*_*i*_ represents the probability that a susceptible individual in simulation age group *i* becomes infected per contact with an infectious individual.

We compare *d* = 1, …, 7 free parameters on the common age grid 0–5, 6–8, 9–11, 12–14, 15–17, 18–19 and 20+ years. Lower resolutions constrain adjacent probabilities to be equal; the three-parameter model uses 0–5, 6–14 and 15+ years. Table 4 lists every resolution. ABC and BO use the same age intervals, calibration bounds and simulator within each comparison. The observation groups remain fixed at 6–8, 9–11, 12–14, 15–17 and 18–19 years.

For a given *θ*, the ABM produces a predicted EBV prevalence profile across *M* = 5 data age groups (which may differ from the simulation age group definitions). We denote the model output by

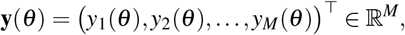

where *y* _*j*_(***θ***) is the simulated prevalence in data age group *j*.

Let 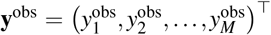 denote the observed mean EBV prevalence rates from the 2009–2010 US National Health and Nutrition Examination Survey (NHANES), reported by Balfour et al.^3^. The five estimates are 50%, 55%, 59%, 69% and 89%, with respective 95% confidence limits [44,56]%, [48,63]%, [54,65]%, [63,75]% and [81,94]%. To account for scale differences and observational uncertainty, we construct transformed and standardized summary discrepancies. Let *Z*_*k*_(*θ*), *k* = 1, …, *M*, denote the *k*-th standardized summary discrepancy between simulated and observed quantities. Here *M* = 5, and

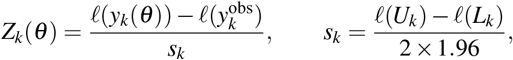

where *l*(*p*) = log(*p/*(1 *− p*)) and *L*_*k*_,*U*_*k*_ are the observed confidence limits. Prevalence is clipped to [10^*−*10^, 1 *−* 10^*−*10^] before taking logits. These standard errors weight the five observations without accounting for covariance between survey groups.

The calibration objective is defined through the Euclidean distance of the standardized summaries, as introduced in Eq. (7)

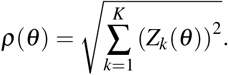

The objective function defined above involves stochastic model outputs due to the inherent randomness of the ABM. We seek parameter values with small expected discrepancy within the calibration bounds,

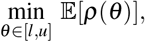

where the expectation is over ABM randomness.

Here *l*_*i*_ and *u*_*i*_ are working calibration bounds. We use [0.002%, 0.006%] for any parameter interval containing ages 0–5 and [0, 0.02%] for all other intervals. Retaining the childhood bounds for pooled intervals makes lower resolutions restrictions of the same seven-interval model. ABC uses a uniform prior on [*l, u*] and BO searches the same bounds. These bounds are calibration choices rather than measured biological limits.

### 4.3 Bayesian Optimization with Expected Quantile Improvement (BO–EQI)

Bayesian Optimization (BO) approaches black-box and noisy objectives by fitting a Gaussian Process (GP) surrogate and using an acquisition function to choose new candidates.

Given a GP posterior with predictive mean *µ*(*θ*) and latent predictive standard deviation *τ*(*θ*), excluding future observation noise, the GP-implied *β* -quantile is

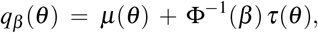

where Φ^*−*1^ is the standard normal quantile function.

At iteration *n*, a simple quantile improvement at *θ* is

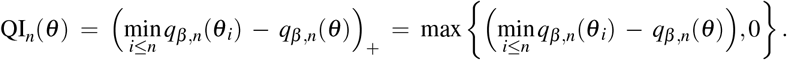

EQI measures the expected improvement of the candidate’s updated predictive quantile over the current best predictive quantile,

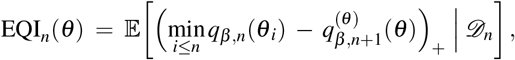

where *D*_*n*_ contains the evaluated parameters and discrepancies, and 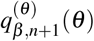 is the future quantile at the candidate after adding its noisy observation.

We initialize the BO procedure with samples generated using Latin Hypercube Sampling^28^ using 20% of the evaluation budget. Each calibration uses 140 initial evaluations out of a total budget of 700. A GP surrogate model is fit to these data, with a single nugget variance estimated to account for observation noise. Optimization is carried out for sequential iterations until the computational budget is exhausted, during which new candidates are selected using the EQI acquisition with *β* = 0.9. The predictive quantile therefore rises with GP predictive uncertainty. The detailed steps of the BO–EQI are presented in Algorithm 2, where *f* = *ρ* and *B* denotes the total evaluation budget. We also refer readers to^27^ for discussion and numerical experiments regarding BO-EQI.

The calibration uses R 4.5.1, `mlrMBO` 1.1.5.1 and `DiceKriging` 1.6.0 with our own EQI implementation. It computes the future quantile 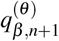 from the latent GP prediction, with the estimated nugget as the observation-noise variance, and was checked against exact Gaussian conditioning and Monte Carlo simulation. To keep the search from re-evaluating points whose prediction is already precise relative to simulation noise, the acquisition is set to zero when the latent predictive standard deviation is below 0.2416, half the root-mean-square standard deviation of *ρ* estimated from 40 simulations at each of seven parameter settings. The returned parameter has the smallest observed discrepancy among evaluated points. Because the smallest of noisy observations is biased downward, its mean discrepancy is assessed using independent validation runs.

#### Algorithm 2 Bayesian Optimization with Expected Quantile Improvement (BO–EQI)

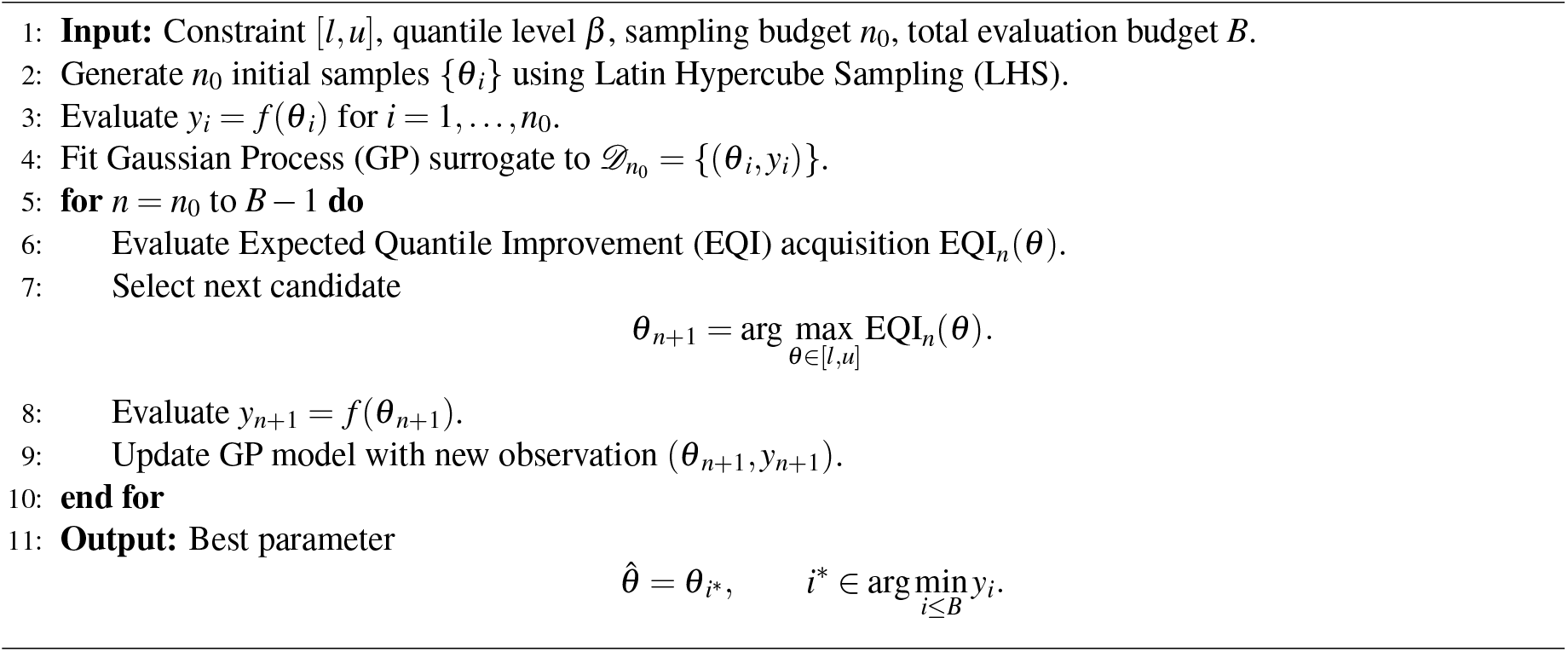

### 4.4 EBV probability of infection calibration: BO-EQI

We first conducted calibration experiments with a single per-contact infection probability parameter. The framework was subsequently extended to include up to 7 age-stratified infection probability parameters, resulting in a higherdimensional calibration problem. We apply both ABC and BO-EQI at each resolution with the same bounds and 700-evaluation budget to compare the effects of parameter resolution.

ABC shares the 140 initial evaluations with BO-EQI and adds 560 Latin hypercube draws from the same uniform prior. It follows the tolerance-selection rule in Section 3, searching fractions from 0.0001 to 0.20 in increments of 0.0001 and requiring at least five accepted draws. The accepted-sample mean is compared with the BO-EQI point estimate. Each fitted point is evaluated with 100 ABM validation runs, independent of calibration, using common seed indices across settings. Table 3 reports their mean discrepancies, Monte Carlo standard errors, and paired 95% intervals for the ABC-minus-BO-EQI difference.

**Table 3.** Matched calibration comparison. Each method uses 700 ABM evaluations at each resolution. Discrepancy means and Monte Carlo standard errors (MCSEs) use 100 validation runs at the BO-EQI point or the ABC accepted-sample mean. The final column gives pointwise paired 95% Monte Carlo intervals for ABC minus BO-EQI; lower discrepancy is better.

| *d* | ABC<br>accepted | ABC mean *ρ*<br>(MCSE) | BO-EQI mean *ρ*<br>(MCSE) | ABC − BO-EQI<br>[95% interval] |
|---|---|---|---|---|
| 1 | 55 | 6.268 (0.054) | 6.175 (0.047) | +0.093 [-0.053, +0.239] |
| 2 | 7 | 6.208 (0.053) | 6.235 (0.052) | -0.027 [-0.167, +0.113] |
| 3 | 5 | 2.086 (0.039) | 1.914 (0.042) | +0.172 [+0.060, +0.284] |
| 4 | 5 | 3.941 (0.055) | 2.484 (0.039) | +1.457 [+1.314, +1.601] |
| 5 | 5 | 9.431 (0.081) | 1.714 (0.033) | +7.717 [+7.545, +7.889] |
| 6 | 5 | 14.838 (0.091) | 2.043 (0.055) | +12.795 [+12.592, +12.998] |
| 7 | 5 | 8.908 (0.081) | 1.799 (0.064) | +7.109 [+6.919, +7.300] |

**Table 4.** Per-contact infection probability estimates by parameter resolution *d* and age, in units of 10^*−*5^. Each cell gives the ABC estimate / BO-EQI estimate on the first line and the marginal 95% credible interval from the posterior. A rule under a group of age columns marks one parameter shared by those ages; all fits use the same five observed groups.

| *d* | 0–5 | 6–8 | 9–11 | 12–14 | 15–17 | 18–19 | 20+ |
|---|---|---|---|---|---|---|---|
| 1 | 2.18 / 2.04<br>[2.02, 2.27] |  |  |  |  |  |  |
| 2 | 2.20 / 2.31<br>[2.01, 2.56] |  |  |  | 2.18 / 1.61<br>[1.00, 3.27] |  |  |
| 3 | 3.56 / 3.94<br>[3.24, 4.72] | 0.740 / 0.582<br>[0.0335, 1.09] |  |  | 3.33 / 3.74<br>[2.13, 5.41] |  |  |
| 4 | 3.36 / 3.41<br>[2.93, 4.58] | 1.42 / 0.889<br>[0.0517, 1.52] |  | 1.30 / 0.0174<br>[0.0120, 2.26] | 2.58 / 5.10<br>[1.81, 5.19] |  |  |
| 5 | 3.25 / 3.97<br>[2.49, 4.38] | 1.93 / 0.107<br>[0.0789, 3.41] | 1.77 / 1.12<br>[0.0178, 1.75] | 3.02 / 0.172<br>[0.0504, 2.08] | 2.73 / 3.59<br>[1.80, 5.13] |  |  |
| 6 | 3.37 / 3.96<br>[2.44, 4.46] | 2.11 / 0.376<br>[0.0538, 3.44] | 2.47 / 0.389<br>[0.0374, 1.90] | 3.11 / 0.00596<br>[0.0446, 2.33] | 4.86 / 4.18<br>[0.228, 4.29] | 7.29 / 8.62<br>[3.13, 19.5] |  |
| 7 | 3.28 / 3.71<br>[2.41, 4.37] | 1.21 / 0.840<br>[0.0676, 3.59] | 1.98 / 0.886<br>[0.0245, 1.78] | 2.57 / 0.940<br>[0.0653, 2.45] | 3.92 / 2.18<br>[0.185, 4.37] | 8.19 / 8.85<br>[2.53, 19.5] | 5.69 / 5.69<br>[0.662, 19.6] |

All calibrations start from the same initial population of 10,000 agents, generated with seed 20260917. They use 28-day steps over 100 years, demographic rebalancing every 0.25 years, and prevalence averaged over outputs stored every 26 steps within ten years of the last stored output. The preceding approximately 90 years are warmup; this duration alone does not establish equilibrium. The simulations are run on one Intel Core i9-13900KF workstation with 32 logical processors and 64 GiB RAM (nominal clock approximately 3.0 GHz).

Table 3 compares the two methods at the same budget of 700 ABM evaluations. For three to seven parameters, the BO-EQI point estimate has a lower mean validation discrepancy than the ABC accepted-sample mean, and the paired 95% interval for the difference excludes zero. For one and two parameters, the two means differ by less than 0.1 and the interval contains zero, so the fits are equivalent at these resolutions.

Table 4 reports the resulting parameter estimates for the data of Epstein-Barr seroprevalence among US children^3^. BO-EQI returns one parameter vector and says nothing about how well the data pin it down, and ABC accepts too few draws at this budget (Table 3) to give intervals, so we estimate parameter uncertainty separately by asking which parameter values are compatible with the survey. A parameter value is compatible to the extent that its simulated prevalence falls within the survey confidence limits. Since *ρ*(*θ*) measures the distance from the observed prevalence in units of survey standard errors, we give each parameter value the weight exp {*−ρ*(*θ*)^2^*/*2{: it equals one for an exact match, about 0.6 when the simulation is one standard error away in a single age group, and about 0.14 at two standard errors. This is the likelihood that follows from treating the five *Z*_*k*_(*θ*) as independent standard normal errors^29^. Combined with the uniform prior *π* on [*l, u*] and averaged over ABM randomness, it gives the posterior

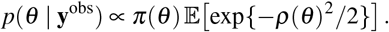

This posterior weights every parameter value continuously, whereas the ABC procedure in Section 3 either accepts or rejects a draw, and it does not use the GP.

The posterior cannot be evaluated on a grid, so we sample it. We draw parameter values, run the ABM once at each, and weight the run by exp {−*ρ*^2^*/*2}. Drawing uniformly from [*l, u*] would spend most runs where the weight is close to zero, so we draw from a distribution *q* concentrated where earlier training simulations found small *ρ* and correct for this preference by dividing each weight by *q*(*θ*_*j*_). The resulting importance weights *w*_*j*_ = *π*(*θ*_*j*_) exp {−*ρ*(*θ*_*j*_)^2^*/*2} */q*(*θ*_*j*_) make the weighted sample represent the posterior above^30^. Here *q* is a mixture of a Student-*t* component fitted to the training simulations and a uniform component over [*l, u*], and it is held fixed during the 145,000 production runs. The intervals in Table 4 are the weighted 2.5% and 97.5% quantiles of these runs. They describe which parameter values the data support and are not centred on either point estimate. At every resolution the weighted sample carries at least as much information as 400 equally weighted draws, and no single draw holds more than 2% of the total weight.

To determine whether the ABC and BO-EQI parameter estimates produce different epidemiological predictions, we compare their five age-specific prevalence outcomes using the 100 common-seed validation runs at each parameter resolution. We regard an absolute mean paired difference smaller than the standard deviation across the 100 validation runs at the BO-EQI estimate as being within the ABM noise floor. A difference comparable to or larger than this standard deviation is at least as large as the variation between individual runs, whereas a paired 95% Monte Carlo interval containing zero indicates that its direction is not stable against simulation noise.

The differences between the ABC and BO-EQI predictions do not generally fall within the ABM noise floor. Across the 35 resolution–outcome comparisons, 27 mean differences are comparable to or larger than the corresponding BO-EQI standard deviation, and only three paired intervals contain zero. In the three-parameter model, four of the five mean prevalence differences exceed this standard deviation; their absolute values range from 1.00 to 3.53 percentage points, and all five intervals exclude zero. In the seven-parameter model, all five differences exceed this standard deviation, ranging from 2.39 to 13.49 percentage points, and all intervals exclude zero.

Figure 8 compares the observed and simulated EBV prevalence by age group across calibration scenarios with increasing parameter resolution. Moving from one to three parameters reduces the mean validation discrepancy from about 6.2 to about 2 for both methods (Table 3). Further splitting produces no monotonic improvement. ABC accepts only five draws at every resolution from three onward, so its accepted-sample mean provides a limited comparison at this budget. These results suggest that moderate parameter stratification is sufficient to reproduce the main prevalence patterns. Additional parameters do not consistently improve the selected fit.

**Figure 8.**
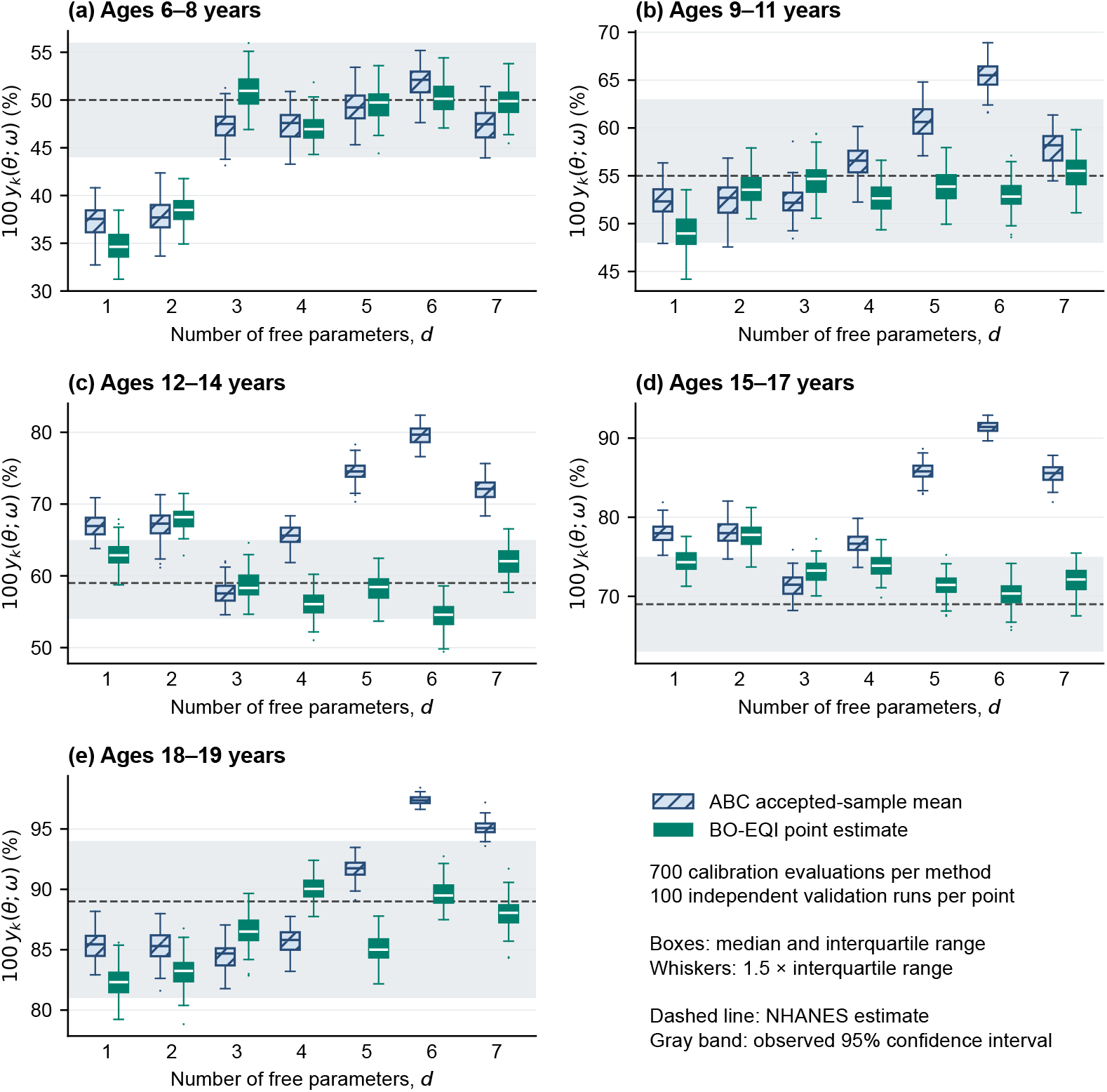
Comparison of observed and simulated EBV prevalence with 100 validation runs per fitted point by age group under each calibration scenario. Navy hatched boxes represent the ABC accepted-sample mean and teal boxes represent the BO-EQI point estimate; the boxes describe simulation variation at fixed parameters. (a) 6–8 years old, (b) 9–11 years old, (c) 12–14 years old, (d) 15–17 years old, (e) 18–19 years old.

Under an equal budget of 700 evaluations, BO–EQI found fitted parameter vectors with lower validated discrepancy than the ABC accepted-sample means at resolutions of three through seven parameters. Rejection ABC yielded only five accepted draws at each of these resolutions, limiting its use for distributional inference at that budget. Conversely, the larger three-parameter ABC run supplied a joint parameter sample that we could propagate through the vaccine scenarios. We obtained marginal intervals for the resolution comparison through a separate importance-sampled posterior, at substantial additional simulation cost and under an explicit discrepancy model. The choice of approach therefore depends on whether the immediate goal is a fitted point or a parameter distribution for downstream analysis.

The vaccination projections in Section 5 propagate the 297 accepted joint parameter vectors from the threeparameter 10,000-simulation ABC calibration in Section 3. Recall that with a separate importance sampling step and under the assumption of Gaussian errors for the prevalence discrepancies as described above, the BO-EQI best-fit point estimate can also be turned into a joint distribution; for simplicity, we use only the ABC-derived posterior here.

## 5. Results

### 5.1 Vaccination Scenarios

As an illustrative application of the calibrated ABM, we examined several hypothetical EBV vaccination scenarios. These were not intended to provide an exhaustive evaluation of EBV vaccine target product profiles, nor to represent any specific vaccine candidates. They were, however, chosen to be broadly comparable to the ones previously explored by Goscé et al.^17^

Given that three parameter strata had been shown sufficient to provide a reasonable fit for our homogeneous population (see Figures 7 and 8), we used the ABC version of the calibration; this provided us with a joint posterior distribution of parameters of size 297. Each scenario was then run as a set of 297 simulations drawn from the posterior. In all scenarios, vaccination was introduced into a population of 10,000 agents that had evolved for 100 years before intervention, thus allowing it to reach a stable epidemiological state. The impact of vaccination was then simulated over the subsequent 100 years and compared with a no-vaccination baseline. Vaccination was handled as a one-time opportunity upon an agent reaching the eligible age, using an uptake of 80% across all scenarios.

We considered three vaccination ages (1, 5, or 11 years), two vaccine efficacies (80% or 100%), and either lifelong protection or waning protection. In the waning scenario, efficacy was assumed to decline exponentially such that, 15 years after vaccination, it remained at 80% of its initial value. Under the exponential decay formulation (Equation 1), this corresponds to a half-life of approximately 46.6 years.

In most scenarios, the vaccine was implemented as being efficacious only against EBV, as in the model of Goscé et al.; no direct efficacy against IM was modeled. However, we also considered a set of scenarios with a vaccine having dual efficacies of 80% against both EBV and IM, both with a 15-year duration of protection. Results are shown in figures Figures 9 to 14. Since IM is an acute (rather than lifelong) condition with highly variable duration, and to align with how real-world IM burden is usually reported, we show the incidence rather than the prevalence for IM.

**Figure 9.**
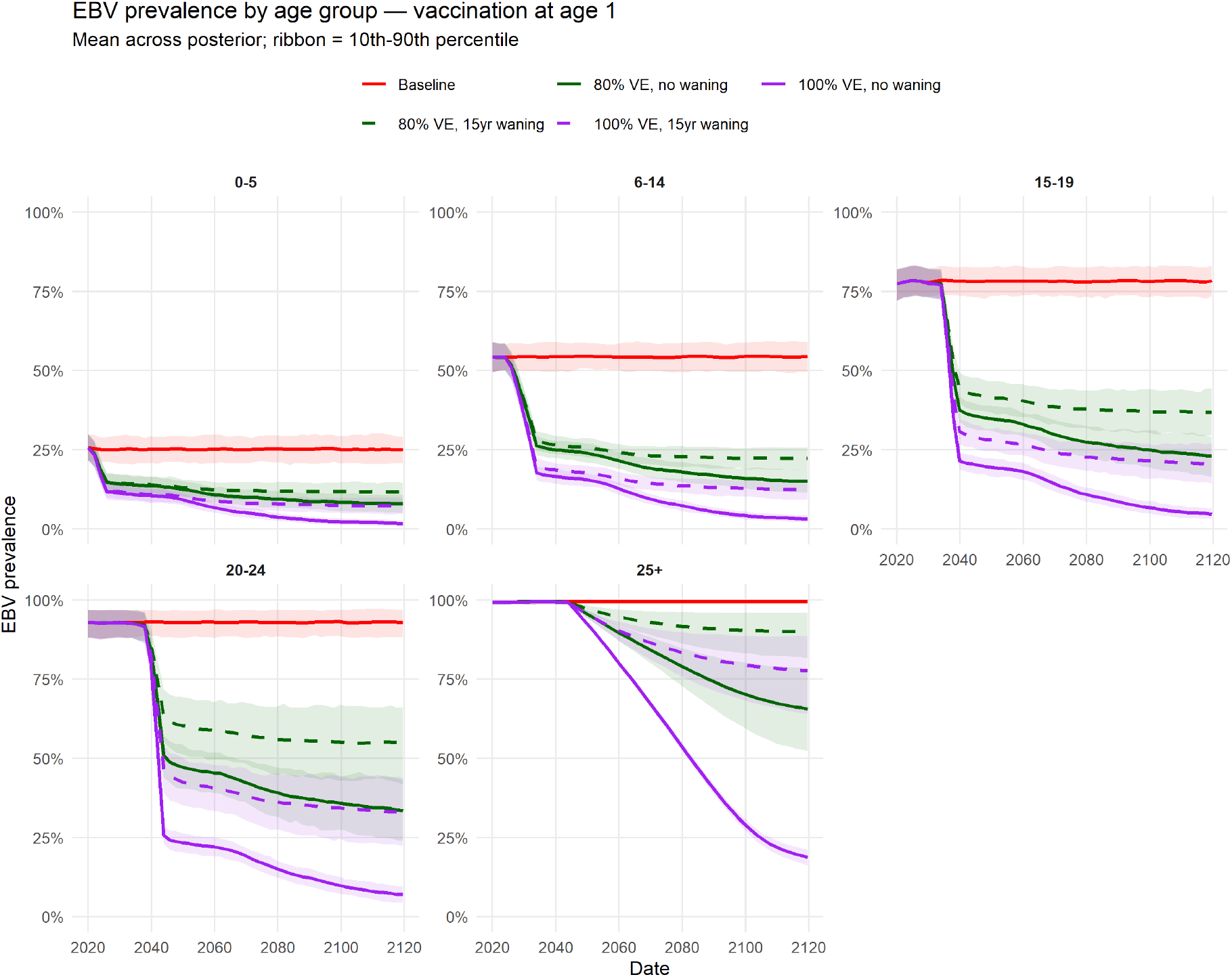
Time evolution of age-stratified EBV prevalence for vaccination at age 1. Scenarios span two values of efficacy (80% and 100%) and protection assumptions (waning, with 80% of initial efficacy remaining after 15 years, or lifelong), together with a no-vaccination baseline. Each trace shows the mean prevalence across simulations with the full posterior. The shaded ribbon is the 10th-90th percentile. Note that since direct efficacy against IM has no effect on EBV transmission, we show the dual-efficacy scenario only in the IM figures, not the EBV figures.

**Figure 10.**
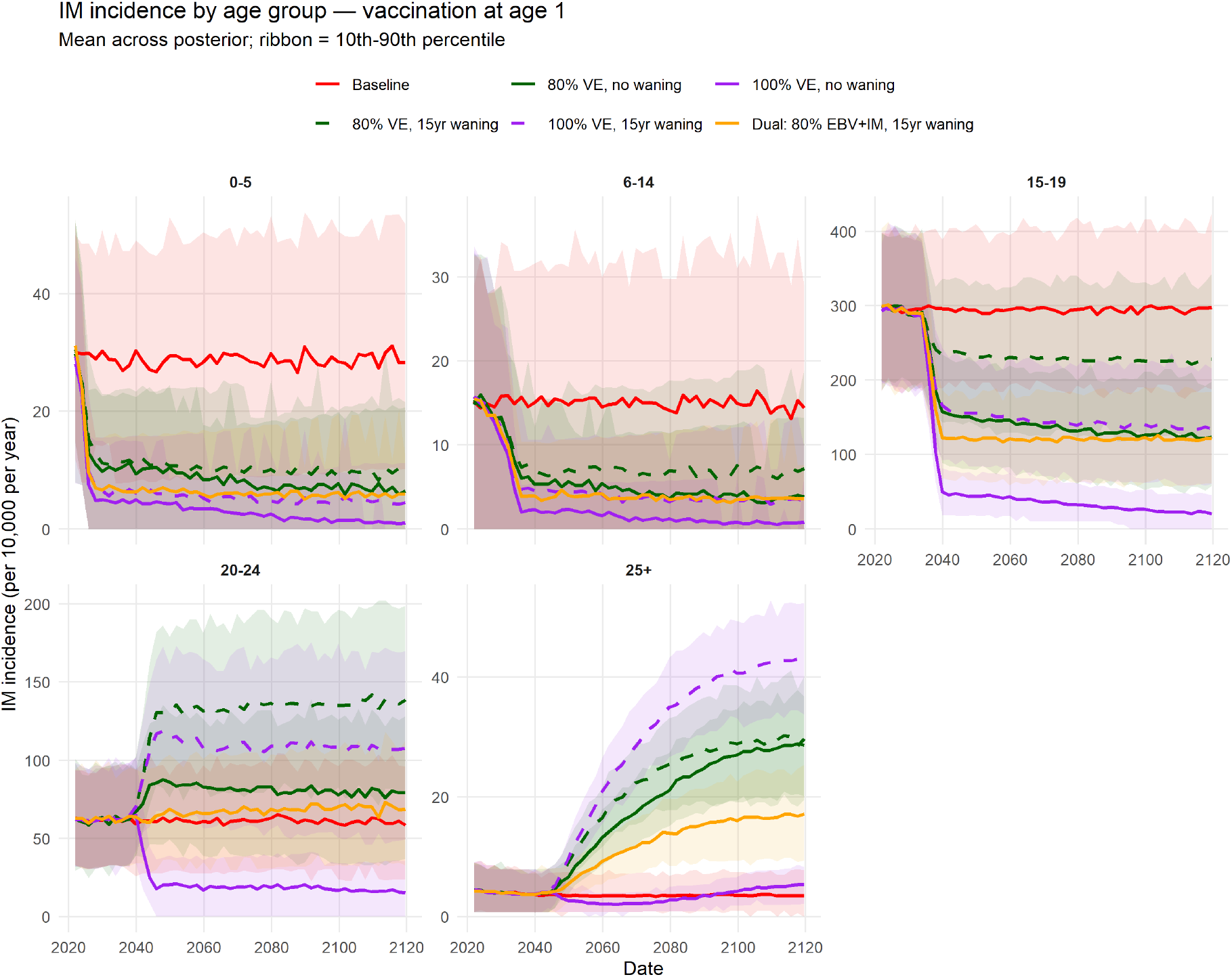
Time evolution of age-stratified IM incidence for vaccination at age 1. Scenarios span two values of efficacy (80% and 100%) and protection assumptions (waning, with 80% of initial efficacy remaining after 15 years, or lifelong), together with a no-vaccination baseline. Each trace shows the mean prevalence across simulations with the full posterior. The shaded ribbon is the 10th-90th percentile.

**Figure 11.**
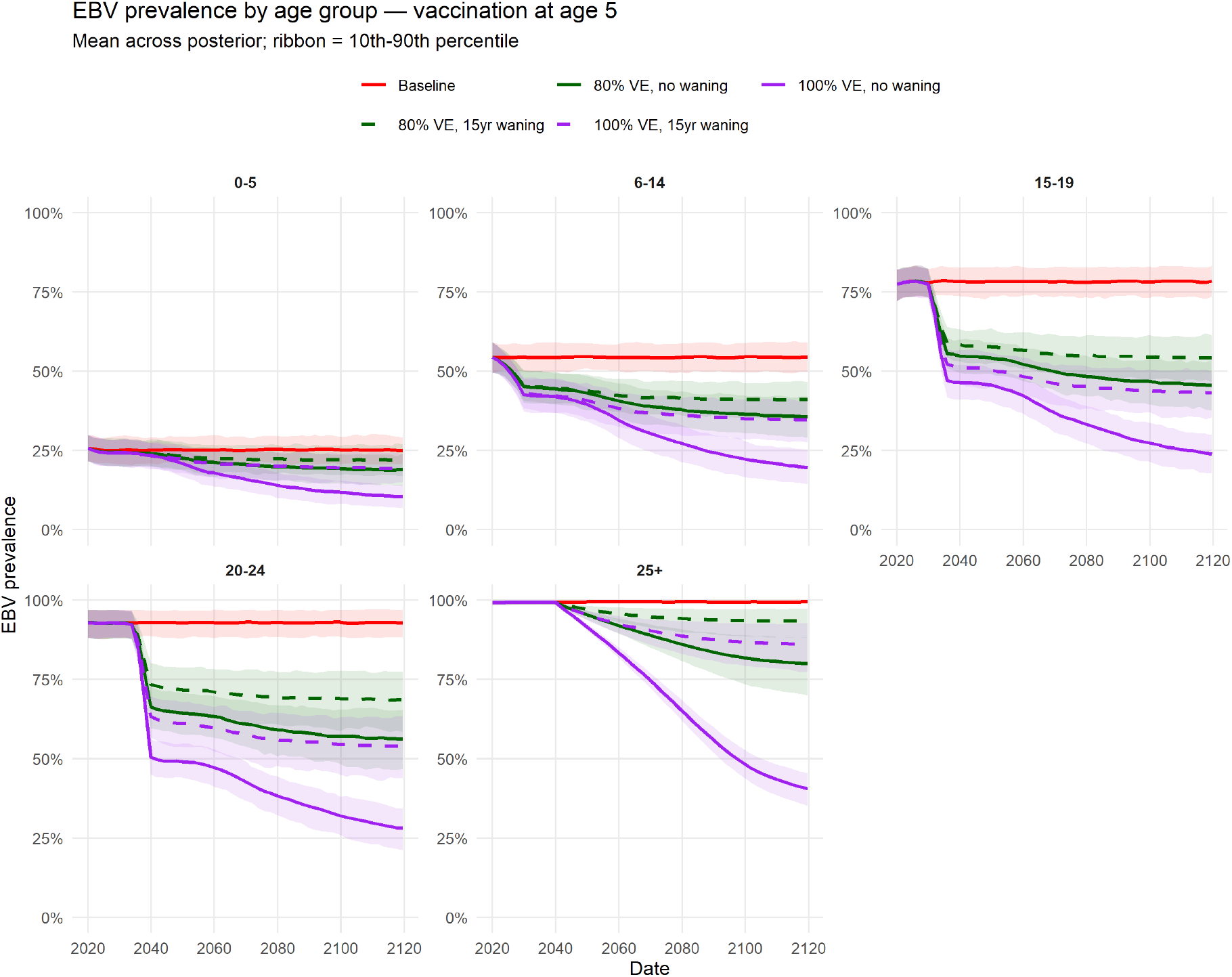
Time evolution of age-stratified EBV prevalence for vaccination at age 5. Scenarios span two values of efficacy (80% and 100%) and protection assumptions (waning, with 80% of initial efficacy remaining after 15 years, or lifelong), together with a no-vaccination baseline. Each trace shows the mean prevalence across simulations with the full posterior. The shaded ribbon is the 10th-90th percentile.

**Figure 12.**
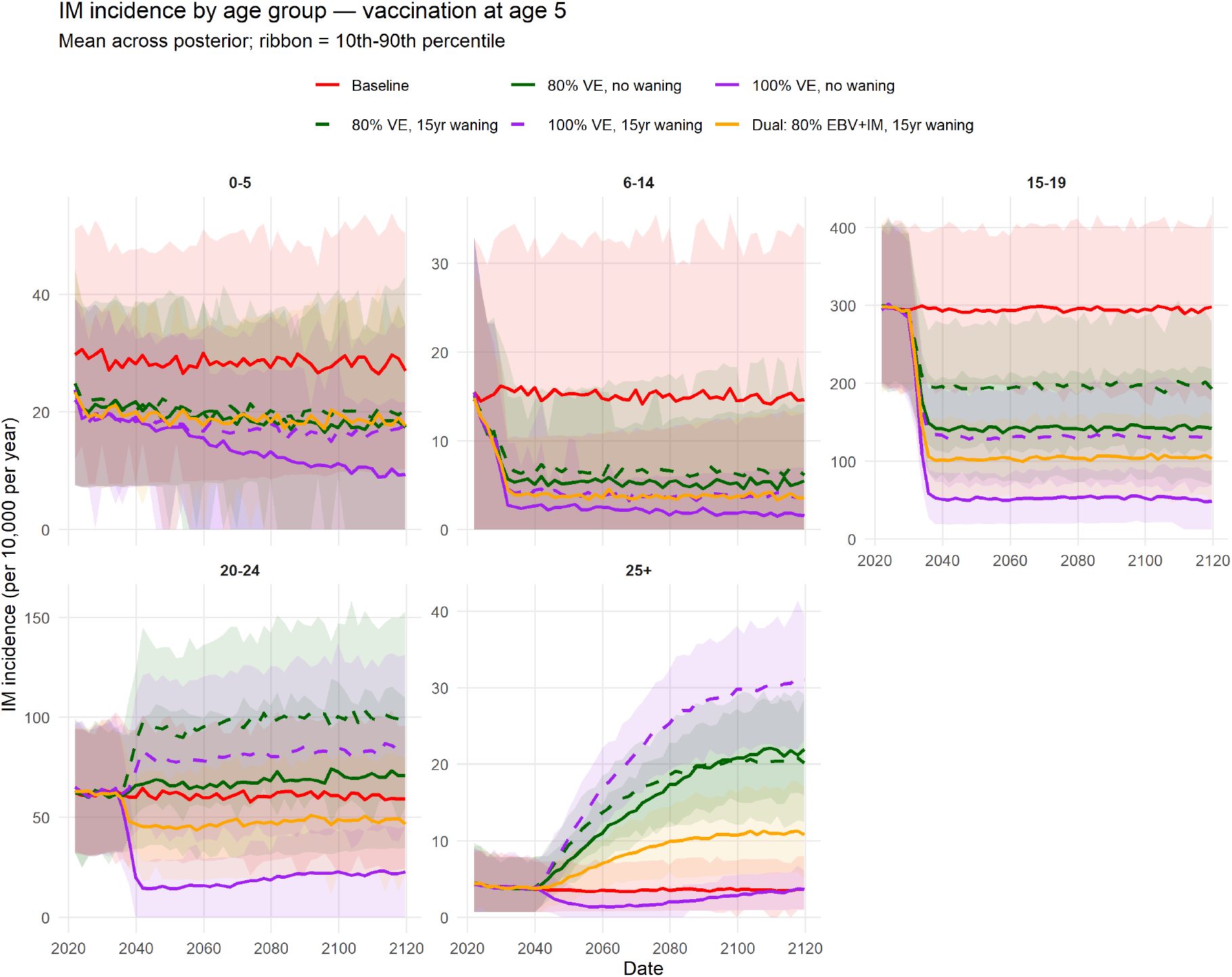
Time evolution of age-stratified IM incidence for vaccination at age 5. Scenarios span two values of efficacy (80% and 100%) and protection assumptions (waning, with 80% of initial efficacy remaining after 15 years, or lifelong), together with a no-vaccination baseline. Each trace shows the mean prevalence across simulations with the full posterior. The shaded ribbon is the 10th-90th percentile.

**Figure 13.**
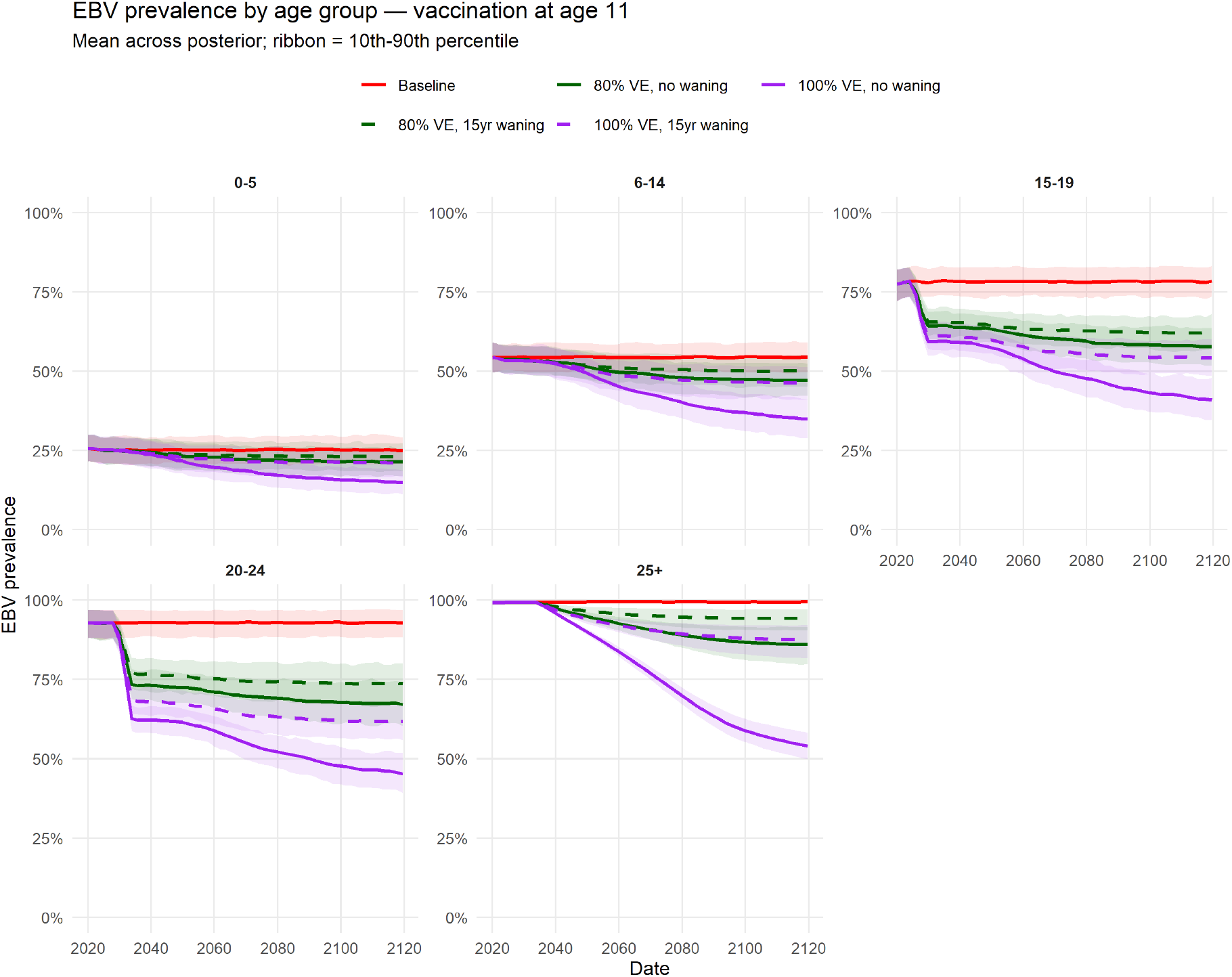
Time evolution of age-stratified EBV prevalence for vaccination at age 11. Scenarios span two values of efficacy (80% and 100%) and protection assumptions (waning, with 80% of initial efficacy remaining after 15 years, or lifelong), together with a no-vaccination baseline. Each trace shows the mean prevalence across simulations with the full posterior. The shaded ribbon is the 10th-90th percentile.

**Figure 14.**
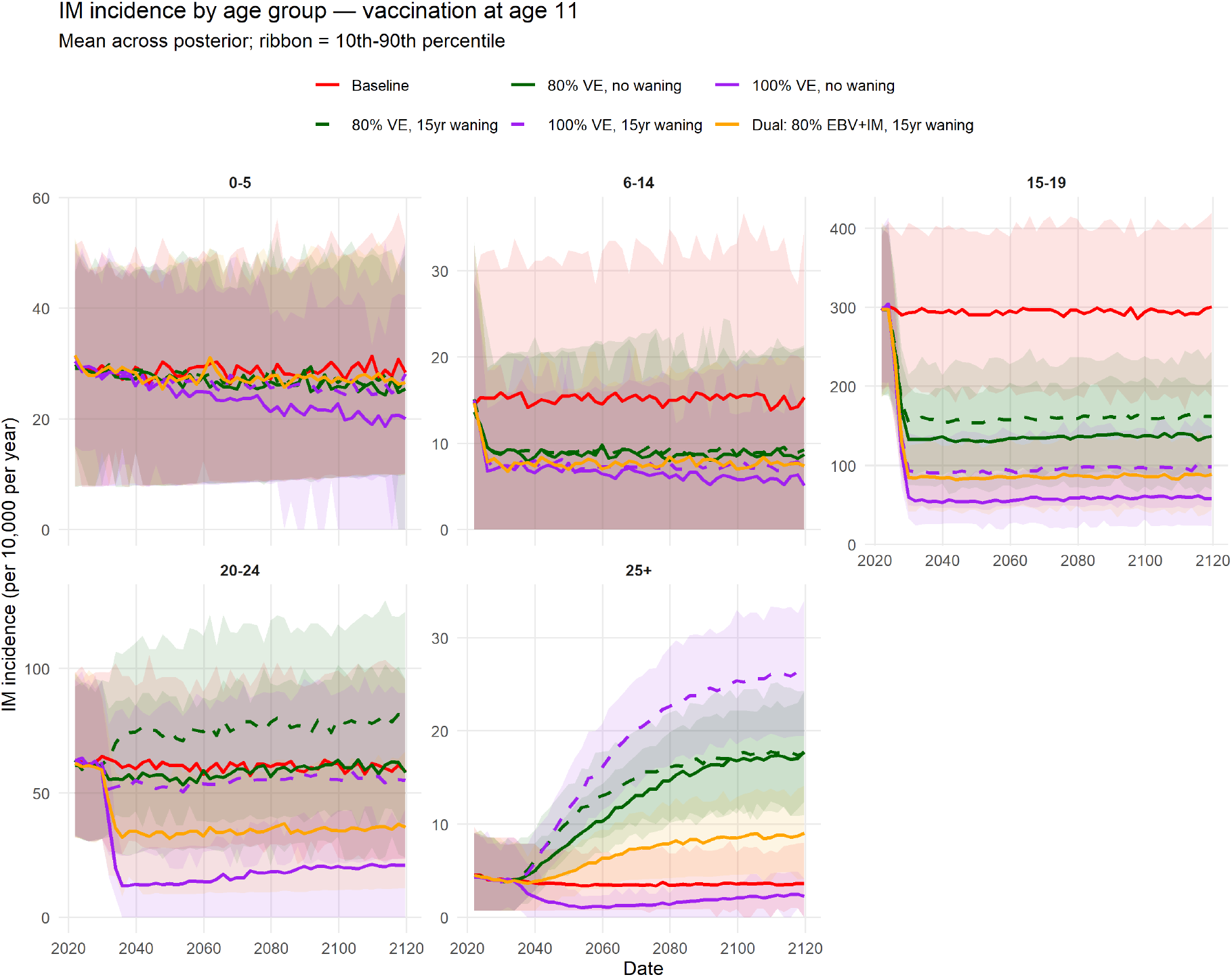
Time evolution of age-stratified IM incidence for vaccination at age 11. Scenarios span two values of efficacy (80% and 100%) and protection assumptions (waning, with 80% of initial efficacy remaining after 15 years, or lifelong), together with a no-vaccination baseline. Each trace shows the mean prevalence across simulations with the full posterior. The shaded ribbon is the 10th-90th percentile.

We observe that the relative impact of vaccine efficacy versus duration changes with age group: Below age 25, increasing efficacy from 80% to 100% achieves roughly the same reduction after 100 years as increasing duration of protection from 15 years to lifetime, whereas in the 25+ group, increase in duration has the greater effect. We also see that for a given efficacy and duration, earlier vaccination achieves greater EBV reduction. In the case of lifetime protection, the reason is simple: Earlier vaccination means fewer years exposed to infection before becoming protected. However, in the case of waning protection, there is a competing effect: Earlier vaccination means earlier waning. Across our scenarios, the former effect is evidently stronger than the latter.

More diverse behavior becomes apparent when we look at vaccine impact on the burden of IM. The “perfect vaccine” scenario (100% efficacy, lifetime protection) remains relatively straightforward; IM incidence is reduced across all age groups below 25, and above 25, where the incidence is already extremely low, it has negligible effect. Furthermore, all scenarios achieved significant reduction in IM incidence in the highest-incidence 12-18 age group, and waning-protection scenarios showed the strongest impact when vaccination took place just before, at age 11, thus minimizing the effect of waning. However, above age 18, the picture becomes more complex. When vaccinating at age 1, all but the perfect vaccine and dual-efficacy scenarios actually increased the IM incidence in the 19-24 age group, by deferring infection to higher ages. Waning further exacerbated this effect by leaving older ages progressively less protected. This effect weakened with increasing vaccination age; when vaccinating at age 11, all but the 80%/15yr scenario reduced incidence relative to the baseline. However, above age 25, all except the perfect vaccine scenario eventually produced elevated IM incidence. Throughout, the dual-efficacy scenario achieved the second-best protection after the perfect vaccine scenario.

### 5.2 Sensitivity of IM Assumption

We examine the sensitivity of our assumption that the probability of developing infectious mononucleosis (IM) following primary EBV infection, *p*_*im*_, remains constant from age 29 onward. The impact of this assumption is most prominent in the IM incidence of the 25+ age group. For vaccination at age one, we consider two alternative assumptions for *p*_*im*_ beyond age 29: a linear decline beginning at age 30 and reaching zero at age 100, and an extreme case in which *p*_*im*_ is set to zero from age 30 onward (Figure 15). Under the original assumption of constant *p*_*im*_, IM incidence increases substantially above the baseline in most vaccination scenarios, except for the non-waning vaccine with 100% efficacy. Introducing a linear decline in *p*_*im*_ reduces the magnitude of these increases, although the overall trends remain similar. In contrast, setting *p*_*im*_ to zero from age 30 onward results in only marginal increases in IM incidence relative to the baseline, with considerably smaller differences between vaccination scenarios. These findings indicate that the projected increase in IM incidence among older individuals is sensitive to the assumed probability of developing IM following primary EBV infection at older ages.

**Figure 15.**
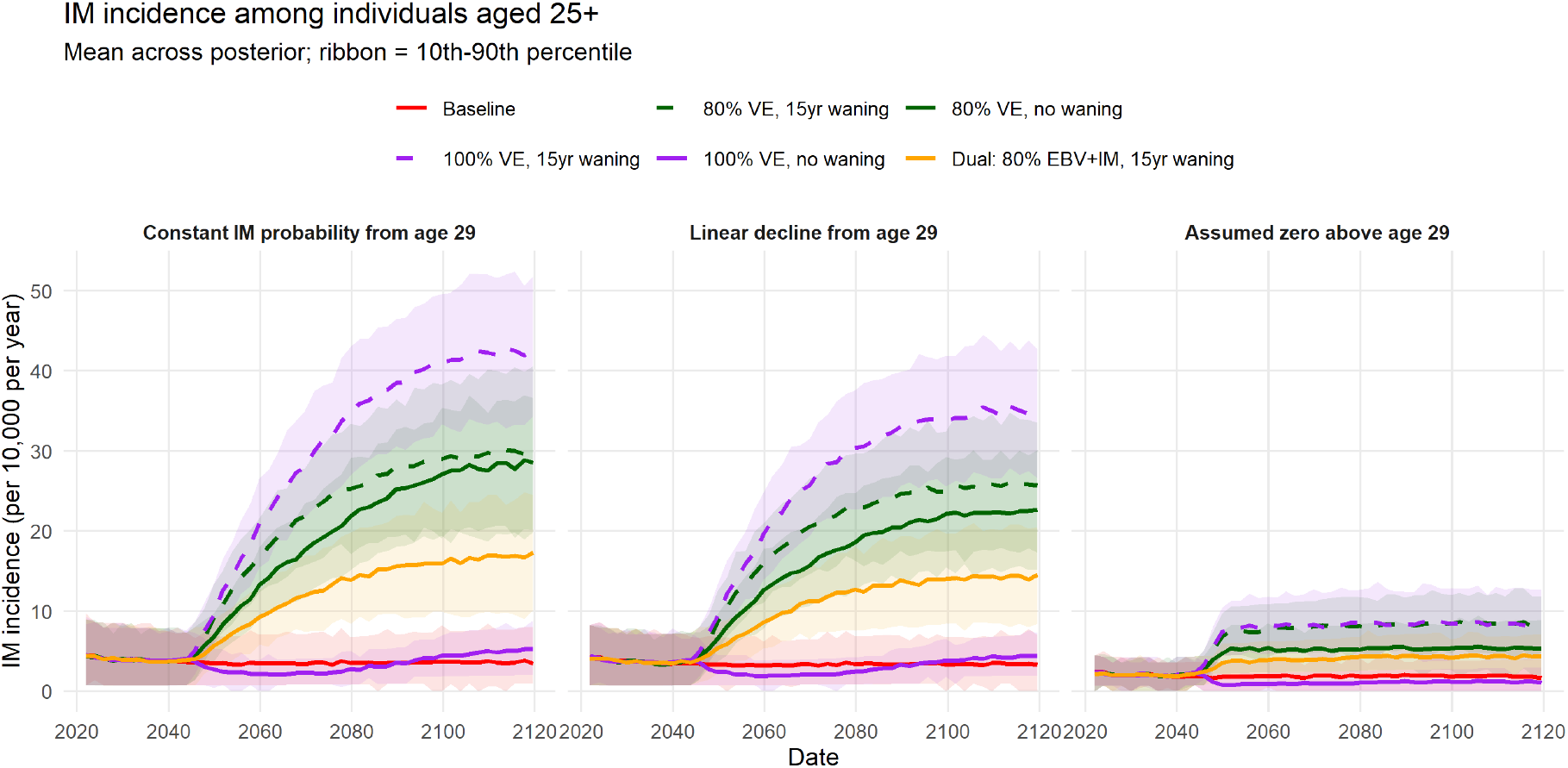
Time evolution of IM incidence in the 25+ age group for vaccination at age 1 under varying assumptions on the probability of developing infectious mononucleosis (IM) following primary EBV infection. Assumptions: (1) original assumption of constant *p*_*im*_ from age 29 onward (2) Linear decline of *p*_*im*_ from age 29 onward (3) *p*_*im*_ is set to zero from age 30 onward. Scenarios span two values of efficacy (80% and 100%) and protection assumptions (waning, with 80% of initial efficacy remaining after 15 years, or lifelong), together with a no-vaccination baseline. Each trace shows the mean prevalence across simulations with the full posterior. The shaded ribbon is the 10th-90th percentile.

### 5.3 Sensitivity of *θ*_1_

There is no well-established nationally representative estimate of EBV prevalence for children aged 0 *−* 5 years in the United States. As a result, the corresponding probability of EBV infection per meaningful contact, *θ*_1_, is not calibrated directly to an age-specific target, but is instead informed by the overall fit of the simulated prevalence profile to the observed age groups. This relies on the assumption that adequately reproducing the downstream age-specific prevalence pattern across all ages provides sufficient information to estimate transmission occurring in the youngest age group. In Figure 16, we divide the baseline scenario into three, based on the input parameter *θ*_1_ in the posterior and investigate the impact of the different range of *θ*_1_ on IM incidence. The scenario labeled “Top 1/3 of baseline” are the baseline results from the highest *θ*_1_ values, “Last 1/3 of baseline” are from the lowest *θ*_1_ values, and “Mid 1/3 of baseline” are the middle range of values. The corresponding *θ*_2_ and *θ*_3_ values of *θ*_1_ remain consistent. This consistency is important to ensure that *θ*_1_, *θ*_2_ and *θ*_3_ are collectively a good fit to the observed EBV prevalence data. In the age range 0-5, all baseline categories overlap. When *θ*_1_ is low, *θ*_2_ compensates with relatively higher values to ensure a suitable fit to the overall observed data. Hence in the age group 6-14, the higher EBV incidence results in higher IM incidence for the lowest *θ*_1_ range, compared to the highest *θ*_1_ range.

**Figure 16.**
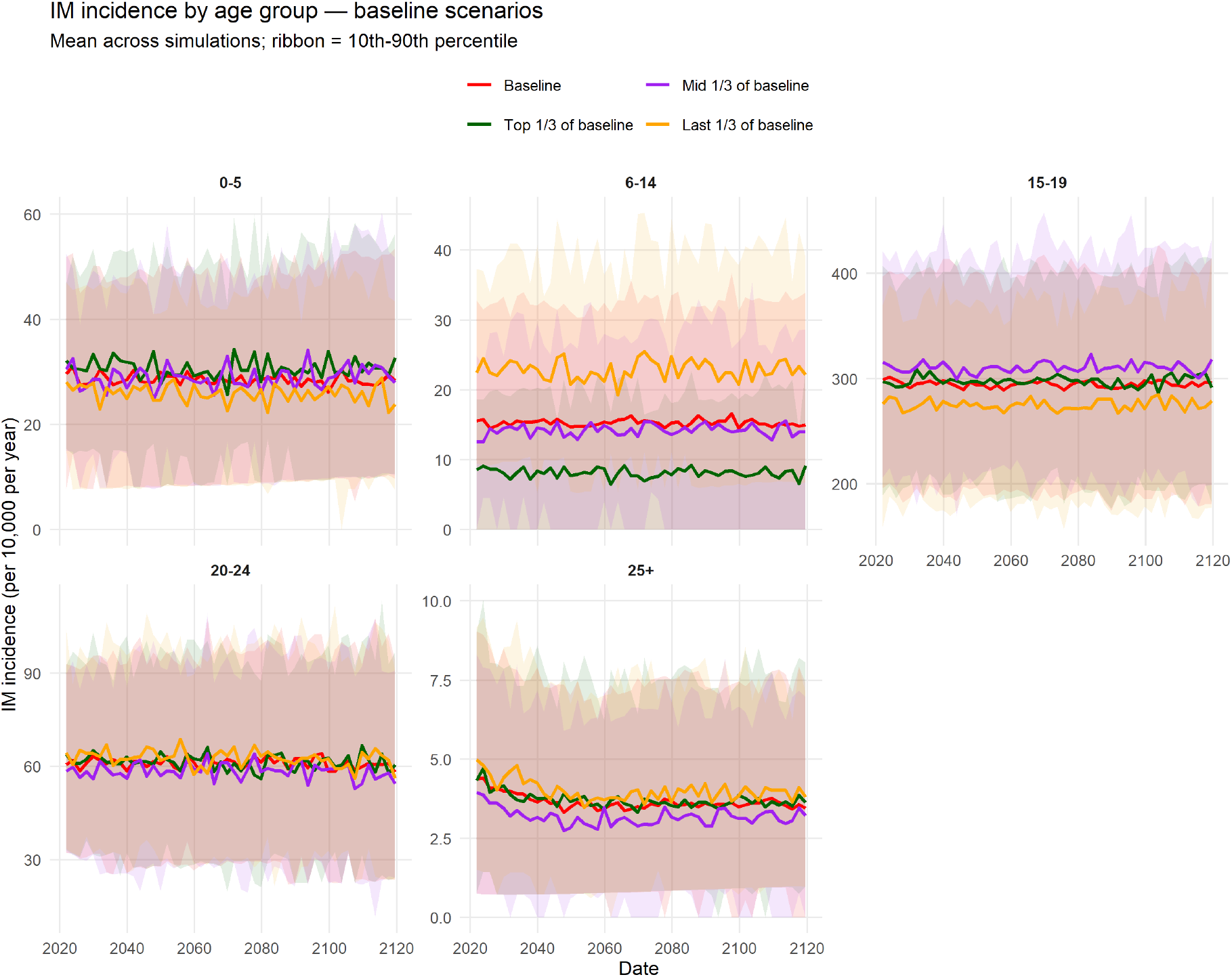
Time evolution of age-stratified IM incidence for baseline scenarios classified based on the input parameter *θ*_1_. There is no vaccination. Each trace shows the mean prevalence across simulations in the subset. The shaded ribbon is the 10th-90th percentile.

## 6. Discussion

In this paper, we developed an agent-based model (ABM) of Epstein–Barr virus (EBV) transmission that tracks infection dynamics within a synthetic population over many decades. By incorporating demographic structure, contact patterns, and age-dependent infection processes, the model provides a flexible framework for examining EBV epidemiology beyond standard compartmental approaches. Calibration to age-specific prevalence data from the United States allowed the simulations to be broadly aligned with available empirical evidence. To our knowledge, this is only the second transmission model of EBV, and the first agent-based model. It is also the first model calibrated to the United States.

A key focus of this work was the comparison of two calibration strategies for estimating the probability of EBV infection per meaningful contact. Using Approximate Bayesian Computation (ABC), we showed how a large ensemble of simulations can be used to obtain a posterior distribution of infection probabilities that provides a good fit to observed prevalence patterns. This likelihood-free approach offers a practical way to account for stochasticity and uncertainty in the model, and naturally yields probability distributions for calibrated parameters. However, there are computational limitations of ABC, particularly as the number of calibrated parameters or the dimensionality of the problem increases, at which point the approach becomes computationally intractable. In the present work, only three transmission parameters required calibration, corresponding to the three age strata in the Balfour et al. data. Had we instead stratified the population by race/ethnicity group as Balfour et al. suggest — given that EBV prevalence varies substantially across such groups^3^ — we would have required nine parameters simultaneously, at which point ABC becomes prohibitively expensive. This represents both the natural use case for BO–EQI and an important direction for future work.

To address the scalability limitations of ABC, we explored Bayesian Optimization with Expected Quantile Improvement (BO–EQI) as an alternative calibration strategy. By formulating calibration as an optimization problem, BO–EQI was able to identify infection probability estimates that reproduce the observed data well while requiring substantially fewer model evaluations. A tradeoff is that BO-EQI yields parameter point estimates rather than probability distributions. At the three-parameter level, BO–EQI and ABC yielded closely comparable results, validating both approaches. At higher dimensionalities, BO–EQI remained tractable where ABC did not, demonstrating its practical value for more biologically detailed parameterizations of heterogeneous populations.

Our vaccination scenario results confirm and extend the findings of Goscé et al.^17^. Consistent with their compartmental model, we find that vaccine durability is at least as important as efficacy in driving reductions in EBV prevalence, and that earlier vaccination achieves greater reduction in EBV burden across the population. We also replicate their observation that vaccination can unintentionally shift the IM burden to older ages. Our results go further, however, in revealing that this is not merely a transient cohort effect. Over Goscé et al.’s 27-year horizon, it was not possible to determine whether elevated IM incidence at older ages would eventually fade away, or whether it represented a durable shift in the endemic equilibrium. Our 100-year projections show clearly that it is the latter: even a century after vaccination introduction, scenarios with waning protection continue to exhibit elevated IM incidence at higher ages relative to the unvaccinated baseline. This persistence reflects the ongoing recycling of susceptible individuals as vaccine protection wanes in each successive cohort.

Our results also illuminate a fundamental vaccine design constraint. The very property that makes infant vaccination most effective for EBV control — intervening before first exposure — simultaneously maximizes the age-deferral effect on IM. Vaccinating at age 1 produces the greatest reduction in EBV prevalence across all age groups, but also produces the greatest increase in IM incidence at higher ages. This tradeoff weakens but does not disappear as vaccination age increases: even at age 11, waning-protection scenarios eventually produce elevated IM incidence above age 25. The implication is that optimizing a vaccine solely for EBV infection prevention, without regard to what happens to deferred infections, risks a net increase in IM morbidity at the population level.

This observation motivated our dual-efficacy scenario, and connects directly to the history of EBV vaccine development. The most clinically advanced candidate to date, the gp350/AS04 subunit vaccine^13^, demonstrated approximately 78% efficacy against IM but no efficacy against EBV infection itself. Conversely, a vaccine with efficacy only against EBV infection reduces transmission and prevalence but does not protect breakthrough-infected individuals from developing IM. Our dual-efficacy scenario is, to our knowledge, the first modelling analysis to quantify what happens when both components act in tandem. The results are clear: across all vaccination ages, the dual-efficacy profile consistently achieves the second-best IM outcomes after the biologically implausible “perfect” vaccine, substantially outperforming EBV-only vaccination at ages 19 and above. This finding argues that dual efficacy against both EBV infection and IM progression should be treated as a core component of future EBV vaccine target product profiles. The recent shift toward mRNA and nanoparticle platforms^15,16^, which offer greater design flexibility than earlier subunit approaches, makes this goal increasingly realistic.

## 7. Limitations and Recommendations

Several limitations of the present analysis warrant acknowledgement. First, the probabilities of developing infectious mononucleosis (IM) following primary EBV infection were estimated from Danish registry data covering ages up to 29 years^20^. Beyond that age, we assume that these probabilities remain constant at their age-29 values. In subsection 5.2, we investigate the impact of declining this probability. The magnitude of the increase in IM incidence in the 25+ age group is lower if the probability is assumed to decrease. However, whether the probability of IM upon primary EBV infection remains high, declines, or approaches zero in middle age is not well established empirically, particularly in the hypothetical setting of vaccination-deferred infection.

The probability of EBV infection per meaningful contact is calibrated to observed data from the US NHANES for ages 6-19. This data is used to calibrate the probabilities across the entire population based on how well the parameters fit to the available observed data. The sensitivity of the parameter representing the 0-5 age group is explored in subsection 5.3. Nationally representative seroprevalence studies in children aged 0-5 years could better constrain early-life transmission parameters. Also, as noted above, the pooling of all race/ethnicity groups into a single calibration may limit results. Balfour et al.^3^ document variations in EBV seroprevalence across racial and ethnic groups in the United States, and a calibration that treated these groups separately would have different transmission parameters (a higher number of parameters) which could provide population dynamics closer to the real world for analysis.

In addition to that, our model does not capture the intermittent and highly variable^31^ nature of EBV shedding by infected individuals. We abstract infectiousness as a constant average value. Other biological complexities that may influence EBV transmission and disease outcomes include vital strain diversity, co-infections and variation in mucosal immunity. Excluding these mechanisms may limit the model’s ability to capture heterogeneity in susceptibility and infectiousness.

Finally, although ABC provides posterior distributions that capture uncertainty, it has high computational cost which increases as the number of calibrated parameters and the population size grows, limiting its practicality for models with higher stratification. BO-EQI addresses this computational challenge and remains tractable at higher dimensions, but at the cost of returning point estimates rather than probability distributions of parameters.

Despite these limitations, this study provides both a methodological framework and substantive epidemiological findings that are relevant to EBV vaccine development. The ABM architecture, with its ability to track complete individual infection histories, is particularly well-suited to the addition of downstream sequelae such as multiple sclerosis and Hodgkin’s lymphoma in future work, outcomes for which the age at primary EBV infection, and its interaction with vaccination, may be of direct clinical importance. For vaccine development and public health planning, model findings could also be translated into clearer target product profiles that distinguish vaccines designed primarily to prevent EBV infection from those that additionally reduce the risk of infectious mononucleosis, including explicit consideration of efficacy, durability of protection, and age at vaccination.

## Data Availability

All data produced in the present study are available upon reasonable request to the authors

## Notes

### Competing Interest Statement

Part of this work was carried out while E.T. was an employee of Sanofi Vaccines. The other authors declare no competing interests.

### Author Declarations

The study used openly available human data that were originally located at https://doi.org/10.1093/infdis/jit321

### Funding

- National Science and Engineering Research Council: 400468, 400871

